# The associations between *Schistosoma mansoni* infection, pre-treatment symptoms, praziquantel side effects, and treatment efficacy in Ugandan school-aged children

**DOI:** 10.1101/2025.05.23.25328199

**Authors:** Huanghehui Yu, Moses Arinaitwe, Adriko Moses, Narcis Kabatereine, Edridah M. Tukahebwe, David W. Oguttu, Aidah Wamboko, Annet Namukuta, Annet Enzaru, Joaquin M. Prada, Alan Fenwick, Joanne P. Webster, Poppy H. L. Lamberton, Jessica Clark

**Affiliations:** School of Biodiversity, One Health and Veterinary Medicine, College of Medical, Veterinary and Life Sciences, University of Glasgow, Glasgow, UK; Section of Paediatric Infectious Disease, Department of Infectious Disease, Imperial College London, London, UK; Vector Borne & NTD Control Division, Plot 15 Bombo Road, P.O Box 1661, Kampala, Uganda; School of Veterinary Medicine, University of Surrey, Guildford, UK; Department of Infectious Disease Epidemiology, Imperial College London, London, UK; Department of Pathobiology and Population Sciences, Royal Veterinary College, Hawkshead, UK

## Abstract

**Background:** Over 240 million people have schistosomiasis. Mass drug administration (MDA) with the anthelmintic praziquantel is the cornerstone of control. Treatment side effects are commonly observed and potentially associated with worms dying in heavily/infected people. However, side effects reportedly reduce MDA uptake, maybe resulting in those most in need refusing repeated treatment. An improved understanding of the association between side effects and infection intensity, pre-treatment health, and drug efficacy, may facilitate improved MDA uptake.

**Methods:** Using latent class analyses and GLMMs we analysed egg and antigen parasitological data and health and side-effects survey data pre- and post-treatment from two primary schools (Bugoto Lake View (LV) and Musubi Church of God (CoG)) in *S. mansoni* high endemicity Ugandan villages to understand whether infection status or intensity were related to 1) pre-treatment symptoms, 2) post-treatment side effects, and 3) whether parasite clearance after treatment was associated with side effects.

**Principal Findings:** At Bugoto LV, abdominal pain, blood-in-stool, and itching/rash were symptoms non-linearly related to pre-treatment infection intensity. Diarrhoea, headache and vomiting were side effects non-linearly associated with infection intensity. At Musubi CoG, blood-in-stool, headache, and pain-when-urinating were non-linearly related to pre-treatment infection intensity. Abdominal pain, diarrhoea, and vomiting were side-effects non-linearly associated with infection intensity. Infection status was not related to pre-treatment symptoms or post-treatment side effects at either school. At both schools, there was no association between infection clearance and side effects.

**Conclusions:** We show no evidence that being infected predisposes someone to side effects, nor that side effects are related to treatment efficacy. The variation between schools in relationships between infection intensity and symptoms or side effects suggest that co-infections and co-occurring sources of morbidity may impact which symptoms and side effects are reported, warranting further investigation, supporting informed discussions with communities.

**Author summary:** Schistosomiasis is a neglected tropical diseases, widely treated by mass drug administration with praziquantel, but side-effects often occur, and are also reported as a reason for low drug treatment uptake. This study examines the impact of praziquantel treatment for schistosomiasis at two schools in Uganda, focusing on the relationships between pre-treatment infection intensity, symptoms, side effects, and treatment effectiveness. Our analysis shows that relationships between symptoms or side effects and infection intensity are not linear, and often were most reported in those who did not have the highest infection intensities. However, the side effects and symptoms reported were not consistent across schools. Furthermore, the occurrence of side effects did not correlate with the probability of successful infection clearance, or with the probability of being infected pre-treatment. Despite some discomforts, praziquantel remains effective in treating schistosomiasis. Our findings highlight the complex nature of praziquantel induced side-effects. Further studies are needed to understand how and why pre-treatment symptoms and/or side effects vary even between demographicaly and geographically similar communities and individuals. This may improve community engagement, in turn improving participation in mass drug administration programs, which is vital to achieving the World Health Organization’s 2030 elimination goals.

## Introduction

Schistosomiasis, is a neglected tropical disease (NTD) with more than 240 million people infected, across 78 countries (1–3). It is prevalent in tropical and sub-tropical regions and disproportionately affects people living in resource-limited settings, with inadequate access to sanitation and safe water. Infection occurs through contact with contaminated freshwater, when the human infective stage of the lifecycle, cercariae, penetrates the skin. In the case of *Schistosoma mansoni*, a major causative agent of intestinal schistosomiasis, adult female and male worms form sexually reproducing pairs and move into the terminal branches of the mesenteric veins, where each pair can produce between 100 and 300 eggs each day (4). Eggs are excreted in the stool and hatch into miracidia once they contact warm freshwater. Once hatched, the micracidia infect snails of the genus Biomphalaria, where they asexually reproduce such that the snails shed thousands of cercariae infectious to humans, completing the life-cycle.

Due to the adult worms being inaccessible and therefore directly unobservable in living people, all diagnostics are a proxy for true infection status. There are two World Health Organization (WHO) recommended diagnostics for *S. mansoni*, which differ in their strengths and weaknesses (3). Kato-Katz thick smears, which count eggs in stool, are highly specific and give an estimate of infection intensity, but egg shedding is highly variable between samples and days, and Kato-Katzs lack sensitivity, often underestimating prevalence, and therefore overestimating drug efficacy (5). The point-of-care circulating cathodic antigen test (POC-CCA) detects antigens that have been regurgitated by feeding worms and are excreted in host urine, and is more sensitive than Kato-Katzs, but lacks specificity and is only semi-quantitative (6). Latent class analyses have been developed to provide greater insight from these imperfect diagnostics, enabling improved estimates of infection prevalence, to guide diagnostic use, and inform on drug efficacy (7–9).

Whilst Kato-Katz and POC-CCAs indicate infection presence and intensity, they do not inform about infection induced morbidity. Eggs that are excreted in the stool are important indicators of ongoing transmission, but a proportion of eggs remain in the body and it is these that cause the majority of the morbidity (10). Acute schistosomiasis is generally associated with a fever and other symptoms collectively known as Katyama Fever (11), but untreated infections often lead to chronic schistosomiasis. This has been associated with impaired cognitive potential, granuloma formation, hyperaemia, anaemia, oedema, colonic ulceration and stunted growth (12–14). Mild chronic infections can cause symptoms such as abdominal pains, blood-in-stool or urine, diarrhoea, dysuria, and dysentery (4,15). In cases of prolonged and heavy infections, more severe illnesses can develop, including hepatosplenomegaly, and it is estimated that schistosomiasis contributes to up to 200,000 deaths a year (16).

Mass drug administration (MDA) with the anthelmintic praziquantel has been the mainstay of control efforts for the last 20 years (17–19). Refering to the distribution of treatment to a target population regardless of the recipient’s infection status, MDA is recommended by the WHO for the control of six NTDs including schistosomiasis (17). The initial aim of MDA was to reduce infection intensities, and associated morbidity, with the indirect aim of reducing transmission (20). Praziquantel is the only existing drug approved for the treatment of schistosomiasis (21,22). A standard dose of 40 mg/kg is recommended, but treatment efficacy can vary greatly (23–28). Additionally, treatment is ineffective against juvenile worms (those less than approximately seven-weeks-old) (29–32), which likely contributes to the rapid resurgence often seen post-treatment (28,33), especially in high endemicity areas where exposure can be constant, such that juvenile worms and new, post-treatment exposures, can be common and rapid. Initial hypotheses suggested that praziquantel exerts its effects through calcium channel interactions in *S. mansoni* (31). More recent findings have provided direct evidence that praziquantel activates a native cation current in the parasite, inducing calcium influx and disrupting its calcium homeostasis (31,32). Recent work has also shown that praziquantel causes damage throughout the tegument of *S. mansoni*, leading to rapid depolarization, muscle contraction, and subsequent death of the adult worms (32). It is hypothesised that a spike in adult worm mortality caused by praziquantel treatment can cause a sudden rush of antigens to be released, which then causes side effects (21,34). Common side effects of praziquantel that have been reported include abdominal pain/stomach discomfort, diarrhoea, dizziness, headache, nausea and vomiting (13). Previous work has shown that between 30% and 60% of patients experience side effects after taking praziquantel, but they are usually mild and disappear within 24 hours (35–37). Side effects can cause patients to refuse treatment in subsequent rounds (36,38), including if the recipient knows they are infected and so thinks the side effects will be more intense (39). However, it is still unclear whether the presence or intensity of symptoms or side effects are connected to pre-treatment infection presence or intensity (40,41). Furthermore, due to the fact that adult worms cannot be directly observed, it remains unclear whether the presence of side effects is indicative of treatment efficacy - i.e., whether more severe side effects reflect greater damage to the worms, leading to more successful clearance, or higher egg reduction rates.

As of 2021, schistosomiasis has been targeted by the WHO for elimination as a public-health problem (EPHP) by 2030 (validated when <1% of school-aged children have a heavy infection intensity, classified as over 400 eggs per gram (epg) of stool using one Kato-Katz for *S. mansoni*). As MDA remains the cornerstone of achieving this ambitious target, it is necessary to understand the interaction between infection prevalence and intensity, symptoms, side effects, and treatment efficacy, so that evidence-based information can be given to communities regarding the risk of side effects and what they might expect as a result of treatment. Here we leverage a latent class analysis framework (7,28) fit to egg- and antigen-based epidemiological diagnostic data as one approach to infer the unobservable, ‘true’, infection status, thereby enabling a more robust estimate of true clearance after treatment. We then use the model ouputs to investigate how pre-treatment symptoms and post-treatment side effects relate to infection dynamics and treatment efficacy. Specifically, we address five main hypotheses: 1) People with *S. mansoni* infections are more likely to report infection-associated symptoms than those who are uninfected; 2) These symptoms are more likely in people with heavier infections in comparison to uninfected people or those with lower infection intensities; 3) People with *S. mansoni* infections are more likely to report side effects when treated with praziquantel than uninfected people; 4) These side effects are more likely in people with heavier infections in comparison to those uninfected or with lower infection intensities when treated with praziquantel; and 5) People reporting more side effects are more likely to have cleared their infections (due to the side effects being potentially caused by damaged *S. mansoni* adult worms – though we do not test this causal hypothesis directly).

## Methodology

### Ethics

Ethical approvals were gained from the Uganda National Council for Science and Technology (Memorandum of Understanding: sections 1.4, 1.5, and 1.6) and the Imperial College Research Ethics Committee (EC NO: 03.36. R&D No: 03/SB/033E). During school committee meetings prior to the start of the study, teachers and community leaders verbally consented to the study happening in their community. Each head teacher gave written permission for the pupils to participate in the study. Before being included in the study, each participant verbally assented and they understood that they could leave the study at any time with no impact on their MDA treatment. This study analysed data that was collected through the National Control Programme that distributes MDA, and all praziquantel treatments were administered by the National Control Programme team who lead the MDA programme in Uganda.

### Study Site and Recruitment

Data were collected from school-aged children at two schools: Bugoto Lake View (LV) and Musubi Church of God (CoG) primary schools, in Mayuge District, Uganda, both based on the shores of Lake Victoria and highly endemic for *S. mansoni*. In 2004, 191 participants were tested (Table 1), with ages ranging from 6 to 12 years old. In Musubi CoG, praziquantel treatment had never previously been administered. In Bugoto LV MDA had started one year previously, in 2003. In both locations, incoming six-year-olds were considered praziquantel naïve as there was no paediatric formulation at the time, and MDA was only administered through schools at this time with six-year-olds having only just started school. All participants were given unique IDs, and sex, height, weight, and age were recorded. Anaemia status was also recorded by using finger prick blood samples and an Hb machine (42) though these specific data are not used here.

**Table 1.** Summary statistics of raw data in Bugoto Lake View (LV) and Musubi Church of God (CoG) in 2004.

|  | **Schools** |  | **Total** |
| --- | --- | --- | --- |
|  | **Bugoto LV** | **Musubi CoG** |  |
| **Sample count (n)** | 123 | 68 | 191 |
| **Sex** |  |  |  |
| Female: | 65 (52.85%) | 34 (50%) | 99 (51.83%) |
| Male: | 58 (47.15%) | 34 (50%) | 92 (48.17%) |
| **Infection prevalence** | 90 (73.17%) | 62 (91.18%) | 152 (79.58%) |
| **Infection intensity (mean ± SD)** | 181.56 ± 258.59 | 433.86 ± 569.07 | 281.31 ± 427.16 |
| **Abdominal pain** |  |  |  |
| Sym: | 71 (57.72%) | 59 (86.76%) | 130 (68.06%) |
| SE: | 68 (55.28%) | 51 (75%) | 119 (62.30%) |
| **Blood in stool** |  |  |  |
| Sym: | 28 (22.76%) | 28 (41.18%) | 56 (29.32%) |
| SE: | 0 (0%) | 0 (0%) | 0 (0%) |
| **Diarrhoea** |  |  |  |
| Sym: | 53 (43.09%) | 55 (80.88%) | 108 (56.54%) |
| SE: | 46 (37.40%) | 33 (48.53%) | 79 (41.36%) |
| **Headache** |  |  |  |
| Sym: | 65 (52.85%) | 59 (86.76%) | 124 (64.92%) |
| SE: | 27 (21.95%) | 23 (33.82%) | 50 (26.18%) |
| **Itching/rash** |  |  |  |
| Sym: | 35 (28.46%) | 37 (54.41%) | 72 (37.70%) |
| SE: | 0 (0%) | 0 (0%) | 0 (0%) |
| **Nausea** |  |  |  |
| Sym: | 41 (33.33%) | 53 (77.94%) | 94 (49.21%) |
| SE: | 0 (0%) | 0 (0%) | 0 (0%) |
| **Urination pain** |  |  |  |
| Sym: | 35 (28.46%) | 29 (42.65%) | 64 (33.51%) |
| SE: | 0 (0%) | 0 (0%) | 0 (0%) |

Sym: pre-treatment symptom; SE: post-treatment side effect.

### Diagnostic Methods

Kato-Katz thick smears were used to quantify *S. mansoni* infection intensities. In both schools, a stool sample was collected once a day for three consecutive days for every participant at each sampling point, and each stool sample was used to process duplicate Kato-Katz thick smears, resulting in six slides per participant per timepoint. Eggs identified as *S. mansoni* were counted under a microscope, averaged across the slides and multiplied by 24 to calculate epg of stool (43). One urine sample was also collected per participant per timepoint and a single POC-CCA test was conducted using the original POC-CCA test with buffer, with results graded as Negative, Trace, +, ++, or +++ depending on the lateral flow assay’s coloured response. After three days of samples had been collected at baseline, all participants were given bread and juice, prior to praziquantel treatment at a dose of 40 mg/kg, regardless of infection status (44) as per the MDA programme for schistosomiasis in Uganda, but using scales for weight rather than the dose pole. At the time, a Kato-Katz test was performed one week after the initial praziquantel treatment to assess treatment response, as it was not yet fully clarified when eggs cleared post treatment to represent drug efficacy. Participants with *S. mansoni* egg counts exceeding 100 epg at this one-week timepoint were administered a second dose. At 1-week post-MDA, 86% of participants in Bugoto and 89% in Musubi had infection intensities above this threshold, indicating that the majority of participants underwent re-treatment. Four weeks after the first treatment the same participants provided three days of duplicate Kato Katz thick smears, and a single POC-CCA test. Here, we do not use the one-week post-treatment data, but use pre-treatment and four-week follow-up data.

### Symptoms and Side-effect Surveys

Pre-treatment, each participant answered a health survey about a range of possible symptoms including abdominal pain, blood in stool, diarrhoea, headache, nausea, pain-when-urinating, and/or rash/itching and if they had ever had them, or had them presently. The survey was provided by the Schistosomiasis Control Initiative (now Unlimit Health) and was used in both intestinal and urinary schistosomiasis areas and therefore included questions on pain-when-urinating. Although not expected to be associated with intestinal schistosomaisis and depsite there being no *Schistoma haematobium* (the cause of urinary schistosomiasis) in the region, pain-when-urinating was reported pre-treatment and therefore kept in the analysis. It has also been significantly associated with reduced health-related quality of life in individuals with and without *S. mansoni* evidencing a complicated relationship between pain-when-urinating and intestinal schistosomaisis that is not well understood (45). Twenty-four hours post-treatment all participants were asked about any side-effects they might have experienced since taking the praziquantel, including abdominal pain, blood-in-stool, diarrhoea, difficulty breathing, feeling dizzy, headache, nausea, pain-when-urinating, itching/rash, face swelling, vomiting, vomiting blood, and/or weakness. Both the symptoms and side-effects were based on self reporting, except if vomiting was observed after treatment, in which case it was noted down and the participant was retreated. If they vomited again they were referred to the health facility and excluded from the study, but this did not affect their access to the MDA of the control programme. Figure S1 summarises the reported symptoms and side effects in relation to parasitological egg counts, and the proportion of side effects and symptoms for each school shown in Figure S2 (Musubi CoG) and Figure S3 (Bugoto LV).

### Data Partitioning & Statistical Tools

The data from Bugoto LV and Musubi CoG were used for the Bayesian Hidden Markov Model (HMM) and data analysis for all hypotheses, and analyses and visualisations were conducted using the open access software R v4.1.2 (46). Data manipulation and visualisations were undertaken with the *tidyverse* package (47). Analysis of residuals for frequentist mixed effects models was conducted using the *DHARMa* package (48). Due to significant differences between the schools in the symptoms and side effects reported, models were run and results reported individually by school.

### Hidden Markov Model (HMM)

To the data from Bugoto LV and Musubi CoG, we fitted an existing HMM framework that combines Kato-Katz and POC-CCA data for each person as imprecise indicators of the latent infection state, to estimate true infection status pre- and post-treatment and subsequently the probability of clearance post-treatment (7,28). Numbers of samples included in the HMM analysis are summarised in Table S1. We used raw repeated egg counts, and POC-CCA scores that were converted such that they ranged from 0 to 4 (equating to Negative (0), Trace (1), + (2), ++ (3), +++ (4)). Further model details are described elsewhere (7,28), but in short, the model predicted the true unobservable egg intensity for each individual drawn from a gamma-distributed population-level mean at baseline for each school, with an autoregressive random walk component for subsequent timepoints. To account for overdispersion in individual egg counts, the likelihood on the Kato-Katz egg counts was drawn from a negative binomial distribution. The likelihood for the POC-CCA data was derived from a logistic function, where the numerator was the maximum possible numerical score (in this case +++ represented as 4), and the real number of the denominator, the true unobservable infection intensity for that individual as estimated by the model. This assumes that as true infection intensity increases, so does the POC-CCA score. In our analysis, we used the POC-CCA scores to reflect a range of infection intensities, rather than the interpretation of results as either positive or negative. We calculated the infection probability for each individual at each timepoint from the estimated latent infection status, over 40,000 iterations with two chains. The population-level clearance and reinfection proportions at each timepoint were drawn from a weakly informative Beta distribution. This estimated proportion was then used to at each timepoint to estimate whether an individual *i* had cleared infection or become reinfected at each timepoint *t* as a 0 or 1, drawn from a Bernouilli distribution. In turn, the clearance probabilities for each individual at each timepoint could be calculated as the proportion of iterations in which the model predicted true clearance for each individual at time point *t* out of all 40,000 iterations. The probability of clearance in relation to each side effect is summarised in Figure S5 for each school. The *runjags* package (49) was used to fit the model.

### Hypotheses 1: Association between infection probability and symptoms

The probability that an individual was infected pre-treatment was used as the explanatory variable, and the symptom reported by participants before treatment (binary variables) was used as the response variable. This was modelled with a generalised linear model with Beta distributed errors by *glmmTMB* package in R (50). A model was run for each symptom (abdominal pain, blood in stool, diarrhoea, headache, itching/rash, nausea, and pain when urinating). The decision to run separate models rather than one model with all symptoms was due to the issue of collinearity. For example, abdominal pain and diarrhoea are not necessarily independent of one another, but equally can occur exclusively. The same logic is applied in the following tests where relevant.

### Hypotheses 2: Association between infection intensity and symptoms

We next asked whether infection intensity could explain the subsequent reporting of symptoms. We randomly sampled 5,000 iterations from the posterior distribution of estimated egg counts. For each iteration, we calculated proportions of egg counts and symptoms and fitted a dose–response model using the *drc* package in R (51). The response variable was the proportion of participants that reported the symptom before treatment. The explanatory variables were the egg count categories, where each category contained 120 epg egg counts (e.g., 1-120, 121-240; 0 eggs as estimated by the model therefore truly no eggs, was considered as a separate category). In Musubi CoG, egg count categories ranged from 0 to 3,000 epg, and, in Bugoto LV, they ranged from 0 to 1680 epg. From the resulting model fits, we extracted the 2.5%, 50%, and 97.5% quantiles of proportion of participants that reported the symptom before treatment to summarise uncertainty. We used the median of all estimated egg counts across the 40,000 iterations from the HMM model to fit a single dose–response mode. This fit represents the average association between egg count and symptom reporting, while the corresponding figure shows the variability around this relationship based on the posterior sampling approach.

### Hypotheses 3: Association between infection probability and side effects

The analysis was identical to hypothesis 1, but with side effects as the response variables.

### Hypotheses 4: Association between infection intensity and side effects

Analysis was identical to hypothesis 2, but the proportion of participants in each egg count category that reported side effects was used instead.

### Hypotheses 5: Association between side effects and the probability of clearance

To assess whether there was an association between post-treatment side effects and the probability that an individual cleared infection after treatment we used a sequence of GLMs with beta error distribution, using the *glmmTMB* package (50), to find whether any of the side effects (abdominal pain, headache, diarrhoea, and vomiting) explained the probability of clearance. The response variable was the probability of clearance obtained from the HMM for each participant, transformed to enable the Beta distribution to accommodate boundary values [0,1] (52). The independent variable was the presence of each side effect after treatment separately. Estimates were obtained for each school.

## Results

### Summary statistics

The data presented in this study encompassed two schools: Bugoto LV and Musubi CoG in Mayuge District, Uganda. The total sample count involved in the Latent Class Analysis (LCA) is 191 participants, with 123 from Bugoto LV and 68 from Musubi CoG (Table 1). Musubi CoG exhibited the highest mean and greatest variance (433.86 ± 569.07) in infection intensities and the highest prevalence (92%) (Figure 1, Table 1).

**Figure 1.**
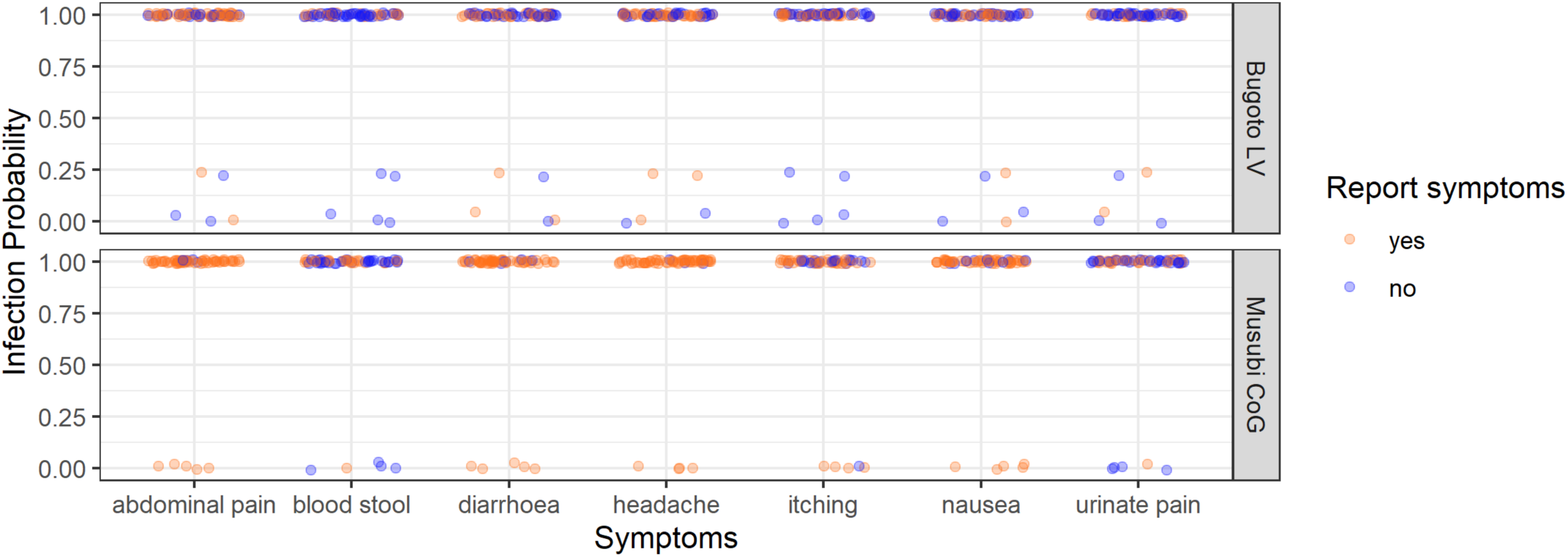
The *Schistosoma mansoni* infection probability (from the latent class analysis) of every participant in Bugoto Lake View (LV) (top graph) and Musubi Church of God (CoG) (bottom graph) primary school who reported (orange), or did not report (blue) pre-treatment symptoms (x-axis) in 2004. Note the jittering of points to prevent them overlaying one another.

The most frequently reported pre-treatment symptom was headache, with a prevalence of 53.57%. This was followed by abdominal pain, reported by 53.02%. Specifically, in Musubi CoG, 86.76% of participants reported headaches, and an equal proportion reported abdominal pain, compared to 52.85% and 57.72% in Bugoto LV, respectively. In terms of post-treatment side effects, abdominal pain was the most commonly reported, with a 45.6% overall prevalence across all three schools. Musubi CoG had the highest prevalence (75%) compared to Bugoto LV (55.28). Other side effects, such as blood-in-stool, itching and/or rashing, nausea, and pain-when-urinating were not reported in Bugoto LV or Musubi CoG.

### Hypotheses 1: Association between infection probability and symptoms

Abdominal pain, blood-in-stool, diarrhoea, itching/rash, nausea, and pain when urinating were reported as symptoms before treatment in both schools. Most participants had a near 100% probability of infection, with only a few participants demonstrating a low probability of infection. This is expected as the shedding of eggs indicates a definitive infection. None of the pre-treatment symptoms were related to the probability of being infected with *S. mansoni* in either of the schools (Figure 1 and Table 2).

**Table 2.**
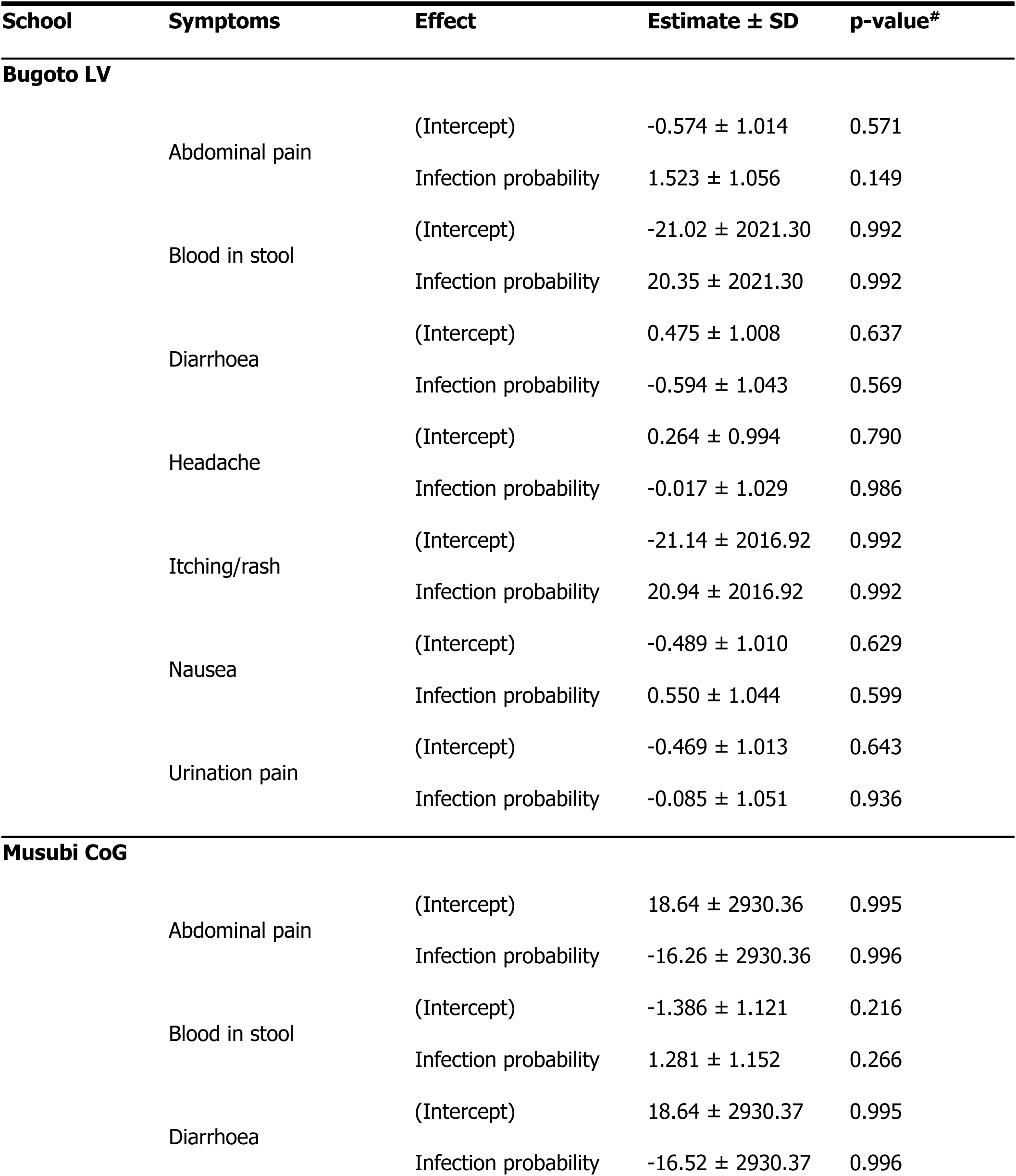
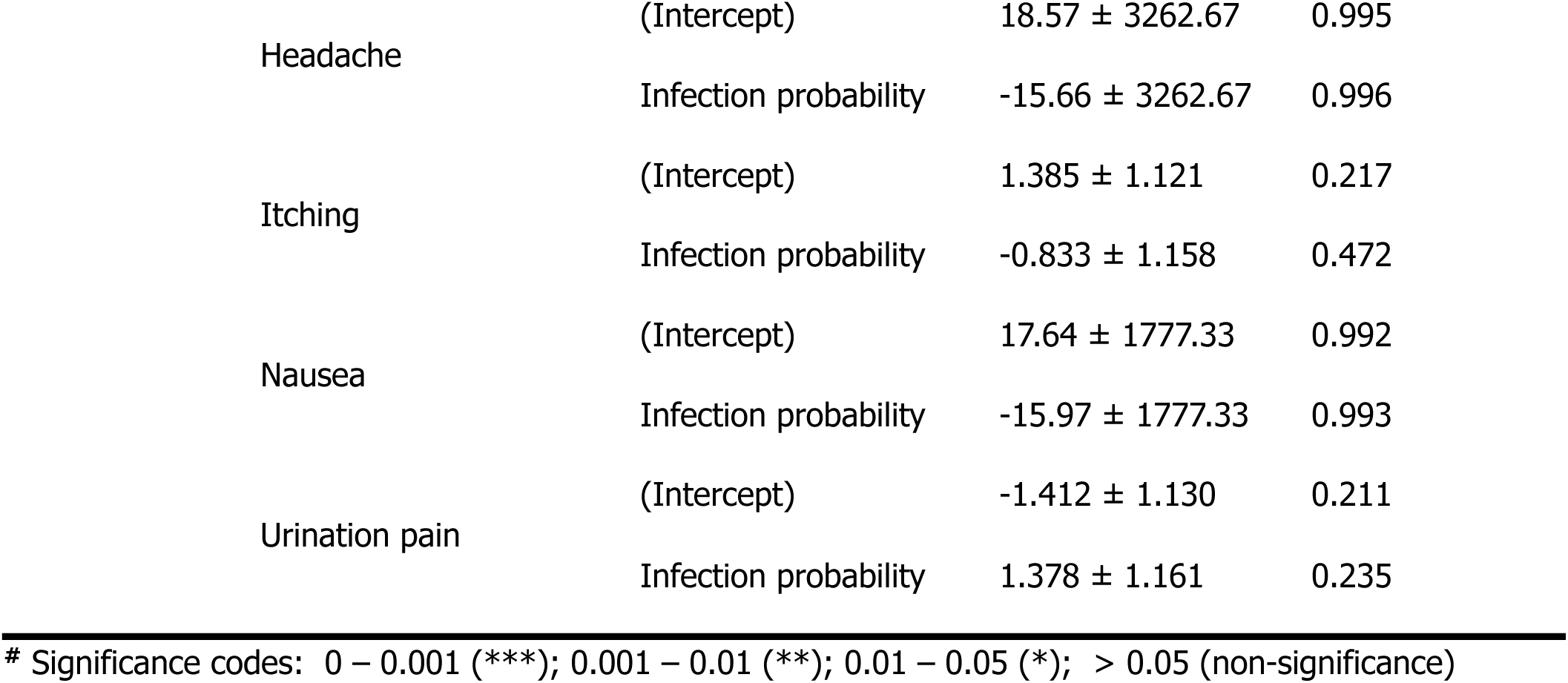
Regression results of the *Beta* model of *Schistosoma mansoni* infection probability (from the latent class analysis) with each symptom as a fixed-effect variable. The intercept was the group of participants who did not report the symptom for each symptom in Bugoto Lake View (LV) and Musubi Church of God (CoG) primary school respectively in 2004.

### Hypotheses 2: Association between infection intensity and symptoms

Abdominal pain, blood in stool, diarrhoea, itching/rash, nausea, and pain when urinating were reported as symptoms before treatment in both schools. The dose–response curves for Bugoto LV (Figure 2, left, pink lines) indicated that increasing *S. mansoni* infection intensity was associated with a significant decline in the reporting of diarrhoea and headache. There is initially almost no relationship between infection intensity and nausea, or pain when urinating, but a significant negative relationship is driven by the decline observed in those with 481-600 epg or more. Alternatively, reports of abdominal pain, blood in stool, and itching display a non-linear relationship with infection intensity, initially showing a positive relationship with increasing infection intensity upto a peak (varies by symptom), before then decreasing with infection intensity (Figure 2, Table 3). Similarly, in Musubi CoG (Figure 2, right, pink lines) reports of abdominal pain, diarrhoea, itching and nausea show little relationship with infection intensity to a point, at which reports declined significantly with rising infection intensity, driven by small samples sizes in the highest egg count categores. The frequency of blood in stool, headache, and pain when urinating also displayed non-linear relationships with infection intensity, with a significant positive relationship peaking before declining once more (Figure 2, Table 3).

**Figure 2.**
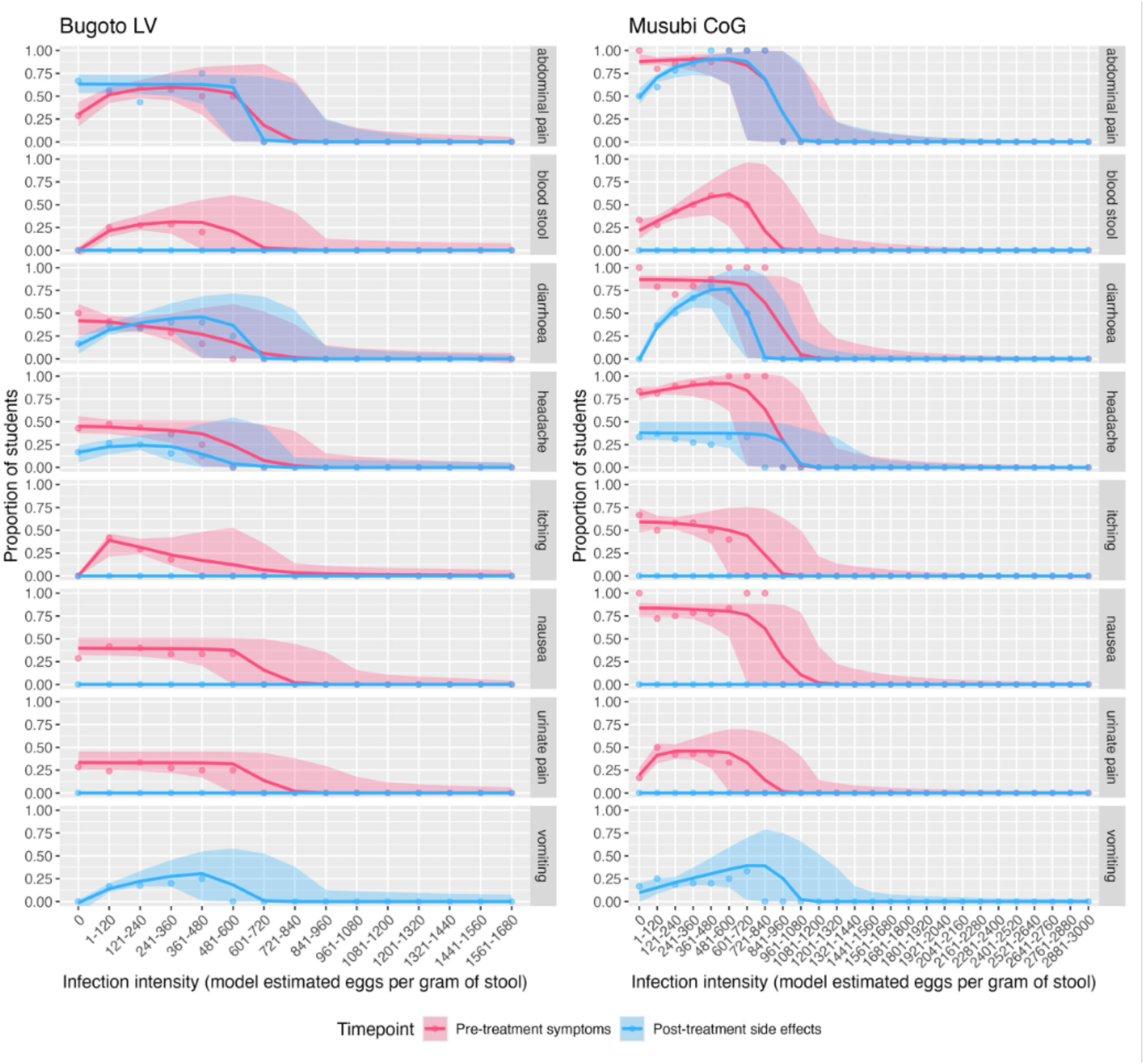
*Schistosoma mansoni* infection intensity dose response curves (from the latent class analysis) for the corresponding symptoms and side effects of *Schistosoma mansoni* infection, including abdominal pain, blood in stool, diarrhoea, headache, itching and rash, nausea, and pain when urinating, for participants from Bugoto Lake View (LV) and Musubi Church of God (CoG).

**Table 3.**
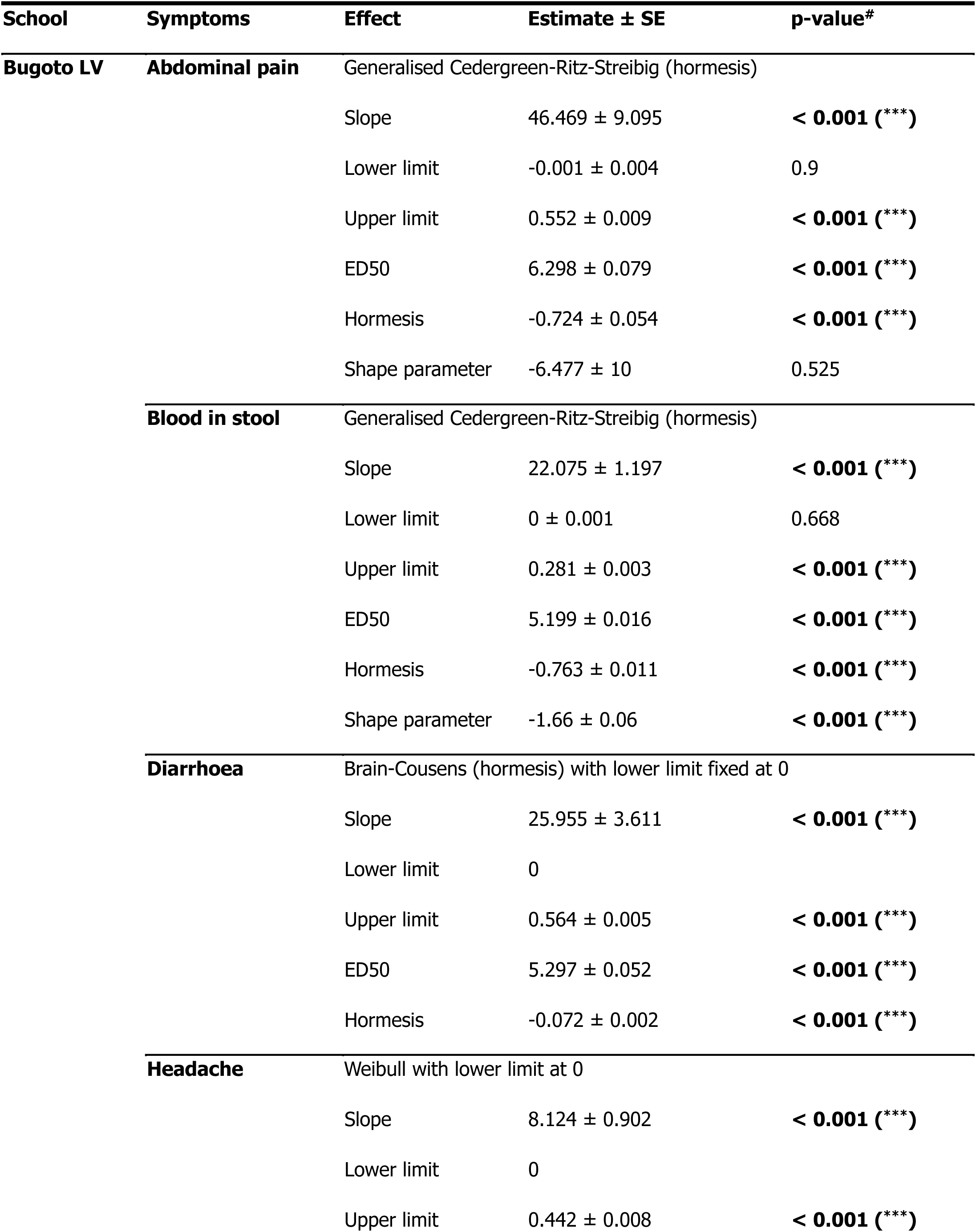
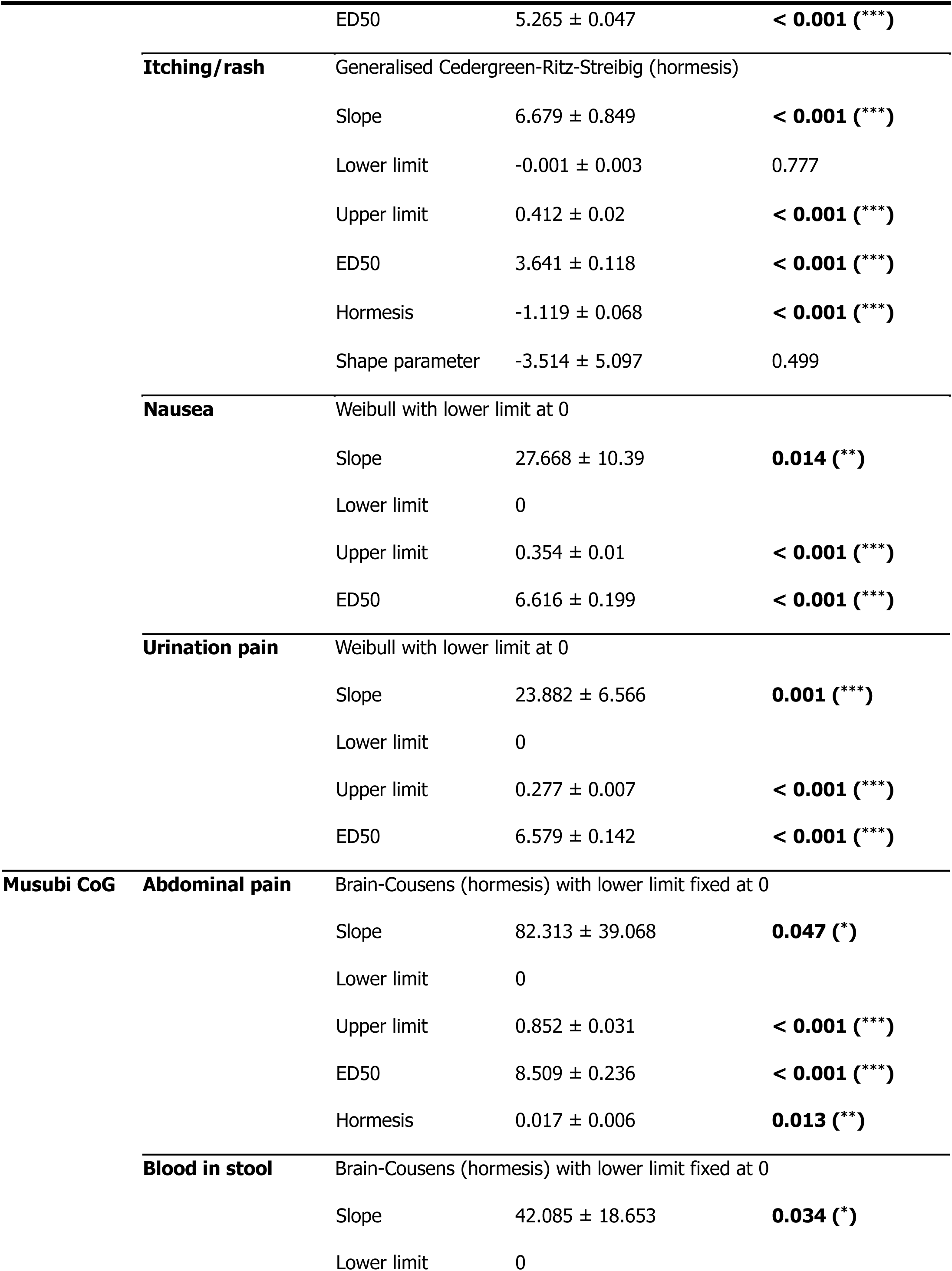
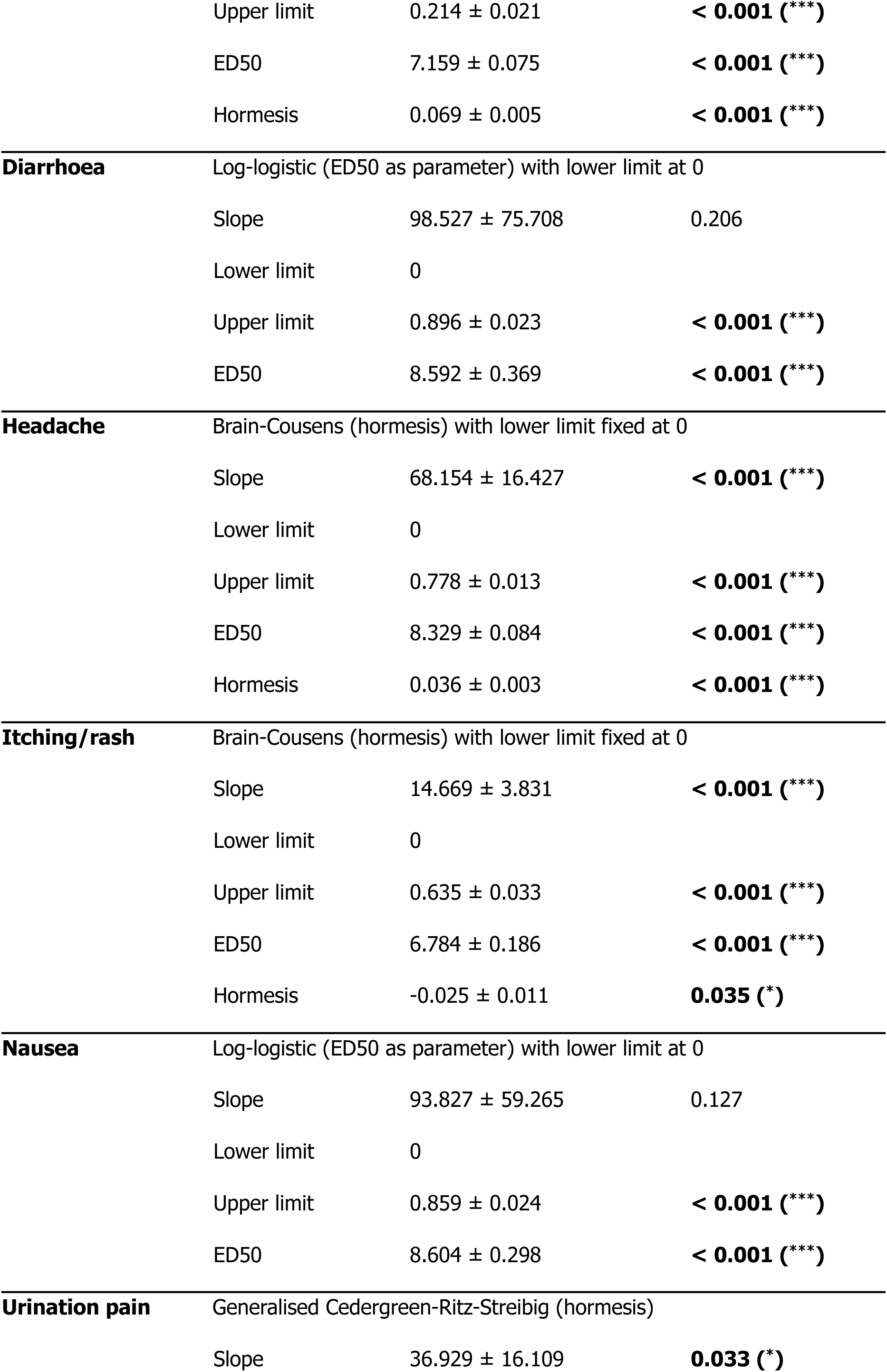
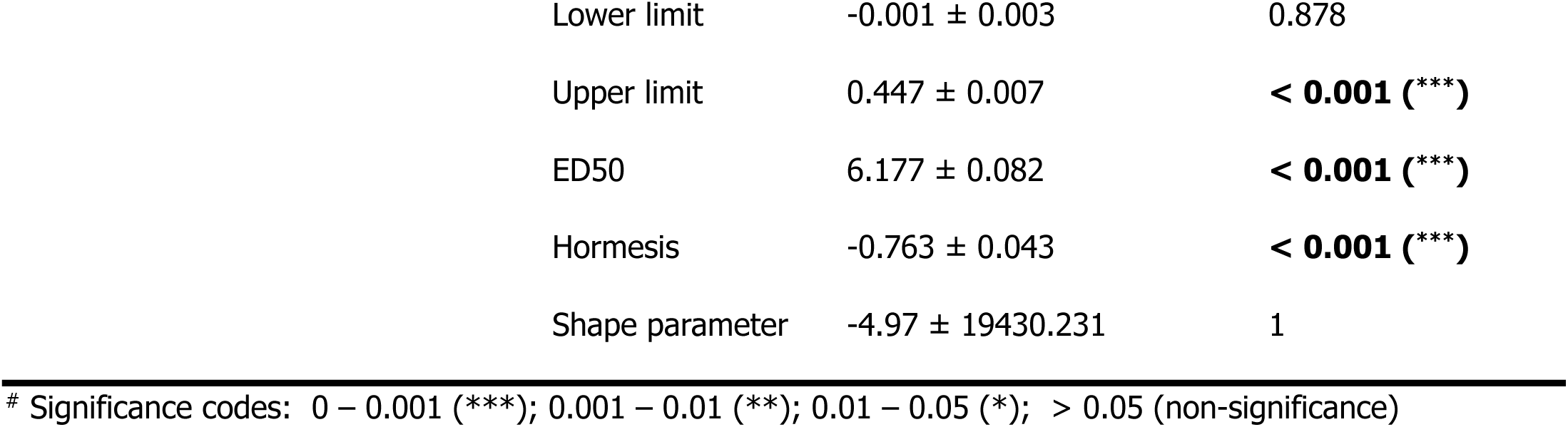
*Schistosoma mansoni* infection intensity dose response results for the pre-treatment symptoms. The intercept was the group of participants did not reporting a symptom or side effect in every symptom in indicated as follows: *** p < 0.001, ** p < 0.01, * p < 0.05, and p ≥ 0.05 is considered non-significant.

| School | Symptoms | Effect | Estimate ± SE | p-value<sup>#</sup> |
| --- | --- | --- | --- | --- |
| Bugoto LV | Abdominal pain | Generalised Cedergreen-Ritz-Streibig (hormesis) |  |  |
| | | Slope | 46.469 ± 9.095 | < **0.001** (***) |
| | | Lower limit | -0.001 ± 0.004 | 0.9 |
| | | Upper limit | 0.552 ± 0.009 | < **0.001** (***) |
| | | ED50 | 6.298 ± 0.079 | < **0.001** (***) |
| | | Hormesis | -0.724 ± 0.054 | < **0.001** (***) |
| | | Shape parameter | -6.477 ± 10 | 0.525 |
|  | Blood in stool | Generalised Cedergreen-Ritz-Streibig (hormesis) |  |  |
| | | Slope | 22.075 ± 1.197 | < **0.001** (***) |
| | | Lower limit | 0 ± 0.001 | 0.668 |
| | | Upper limit | 0.281 ± 0.003 | < **0.001** (***) |
| | | ED50 | 5.199 ± 0.016 | < **0.001** (***) |
| | | Hormesis | -0.763 ± 0.011 | < **0.001** (***) |
| | | Shape parameter | -1.66 ± 0.06 | < **0.001** (***) |
|  | Diarrhoea | Brain-Cousens (hormesis) with lower limit fixed at 0 |  |  |
| | | Slope | 25.955 ± 3.611 | < **0.001** (***) |
|  |  | Lower limit | 0 |  |
| | | Upper limit | 0.564 ± 0.005 | < **0.001** (***) |
| | | ED50 | 5.297 ± 0.052 | < **0.001** (***) |
| | | Hormesis | -0.072 ± 0.002 | < **0.001** (***) |
|  | Headache | Weibull with lower limit at 0 |  |  |
| | | Slope | 8.124 ± 0.902 | < **0.001** (***) |
|  |  | Lower limit | 0 |  |
| | | Upper limit | 0.442 ± 0.008 | < **0.001** (***) |
|  |  | ED50 | 5.265 ± 0.047 | < **0.001** (***) |
|  | **Itching/rash** | Generalised Cedergreen-Ritz-Streibig (hormesis) |  |  |
|  |  | Slope | 6.679 ± 0.849 | < **0.001** (***) |
|  |  | Lower limit | -0.001 ± 0.003 | 0.777 |
|  |  | Upper limit | 0.412 ± 0.02 | < **0.001** (***) |
|  |  | ED50 | 3.641 ± 0.118 | < **0.001** (***) |
|  |  | Hormesis | -1.119 ± 0.068 | < **0.001** (***) |
|  |  | Shape parameter | -3.514 ± 5.097 | 0.499 |
|  | **Nausea** | Weibull with lower limit at 0 |  |  |
|  |  | Slope | 27.668 ± 10.39 | **0.014** (**) |
|  |  | Lower limit | 0 |  |
|  |  | Upper limit | 0.354 ± 0.01 | < **0.001** (***) |
|  |  | ED50 | 6.616 ± 0.199 | < **0.001** (***) |
|  | **Urination pain** | Weibull with lower limit at 0 |  |  |
|  |  | Slope | 23.882 ± 6.566 | **0.001** (***) |
|  |  | Lower limit | 0 |  |
|  |  | Upper limit | 0.277 ± 0.007 | < **0.001** (***) |
|  |  | ED50 | 6.579 ± 0.142 | < **0.001** (***) |
| **Musubi CoG** | **Abdominal pain** | Brain-Cousens (hormesis) with lower limit fixed at 0 |  |  |
|  |  | Slope | 82.313 ± 39.068 | **0.047** (*) |
|  |  | Lower limit | 0 |  |
|  |  | Upper limit | 0.852 ± 0.031 | < **0.001** (***) |
|  |  | ED50 | 8.509 ± 0.236 | < **0.001** (***) |
|  |  | Hormesis | 0.017 ± 0.006 | **0.013** (**) |
|  | **Blood in stool** | Brain-Cousens (hormesis) with lower limit fixed at 0 |  |  |
|  |  | Slope | 42.085 ± 18.653 | **0.034** (*) |
|  |  | Lower limit | 0 |  |

|  |  |  |  |
| --- | --- | --- | --- |
|  | Upper limit | 0.214 ± 0.021 | < **0.001** (***) |
|  | ED50 | 7.159 ± 0.075 | < **0.001** (***) |
|  | Hormesis | 0.069 ± 0.005 | < **0.001** (***) |
| **Diarrhoea** | Log-logistic (ED50 as parameter) with lower limit at 0 |  |  |
|  | Slope | 98.527 ± 75.708 | 0.206 |
|  | Lower limit | 0 |  |
|  | Upper limit | 0.896 ± 0.023 | < **0.001** (***) |
|  | ED50 | 8.592 ± 0.369 | < **0.001** (***) |
| **Headache** | Brain-Cousens (hormesis) with lower limit fixed at 0 |  |  |
|  | Slope | 68.154 ± 16.427 | < **0.001** (***) |
|  | Lower limit | 0 |  |
|  | Upper limit | 0.778 ± 0.013 | < **0.001** (***) |
|  | ED50 | 8.329 ± 0.084 | < **0.001** (***) |
|  | Hormesis | 0.036 ± 0.003 | < **0.001** (***) |
| **Itching/rash** | Brain-Cousens (hormesis) with lower limit fixed at 0 |  |  |
|  | Slope | 14.669 ± 3.831 | < **0.001** (***) |
|  | Lower limit | 0 |  |
|  | Upper limit | 0.635 ± 0.033 | < **0.001** (***) |
|  | ED50 | 6.784 ± 0.186 | < **0.001** (***) |
|  | Hormesis | -0.025 ± 0.011 | **0.035** (*) |
| **Nausea** | Log-logistic (ED50 as parameter) with lower limit at 0 |  |  |
|  | Slope | 93.827 ± 59.265 | 0.127 |
|  | Lower limit | 0 |  |
|  | Upper limit | 0.859 ± 0.024 | < **0.001** (***) |
|  | ED50 | 8.604 ± 0.298 | < **0.001** (***) |
| **Urination pain** | Generalised Cedergreen-Ritz-Streibig (hormesis) |  |  |
|  | Slope | 36.929 ± 16.109 | **0.033** (*) |

|  |  |  |
| --- | --- | --- |
| Lower limit | -0.001 ± 0.003 | 0.878 |
| Upper limit | 0.447 ± 0.007 | < **0.001** (***) |
| ED50 | 6.177 ± 0.082 | < **0.001** (***) |
| Hormesis | -0.763 ± 0.043 | < **0.001** (***) |
| Shape parameter | -4.97 ± 19430.231 | 1 |

### Hypotheses 3: Association between infection probability and side effects

After treatment, there was no statistically significant association between *S. mansoni* pre-treatment infection probability and the presence of any side effects in both schools (Figure 3, Table 4). However, no participants reported blood-in-stool, itching and rash, nausea, and pain-when-urinating as post-treatment side effects, whereas some participants did report them as symptoms (Figure 1).

**Figure 3.**
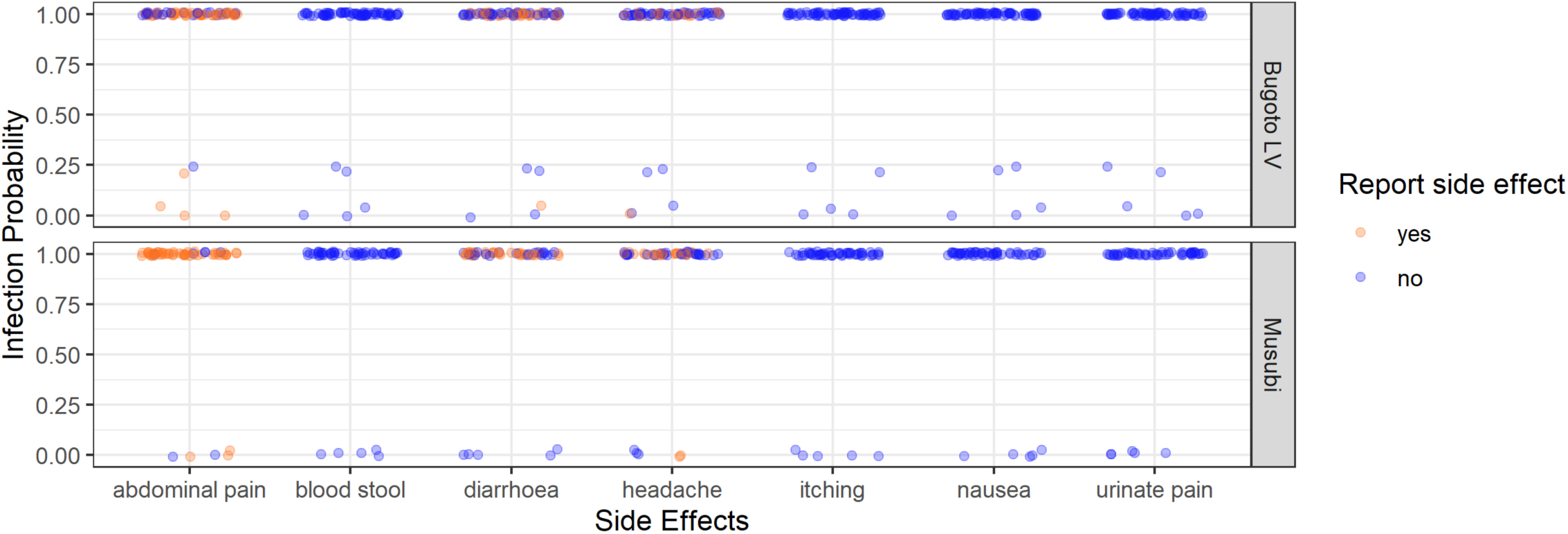
The *Schistosoma mansoni* infection probability (from the latent class analysis) of every participant in Bugoto Lake View (LV) (top graph) and Musubi Church of God (CoG) (bottom graph) primary school who reported (orange), or did not report (blue) post-treatment side effects (x-axis) in 2004. Note the jittering of points to prevent them overlaying one another.

**Table 4.** Regression results of the beta model of *Schistosoma mansoni* infection probability (from the latent class analysis) with each side effect as a fixed-effect variable. The intercept was the group of participants who did not report the side effect for each side effect in Bugoto Lake View (LV) and Musubi Church of God (CoG) primary school respectively in 2004.

| School | Side Effects | Effect | Estimate ± SD | p-value<sup>#</sup> |
| --- | --- | --- | --- | --- |
| Bugoto LV |  |  |  |  |
| | Abdominal pain | (Intercept) | 1.669 ± 1.317 | 0.205 |
| | | Infection probability | -0.9425 ± 1.346 | 0.484 |
| | Diarrhoea | (Intercept) | -1.415 ± 1.209 | 0.242 |
| | | Infection probability | 1.213 ± 1.237 | 0.327 |
| | Headache | (Intercept) | -1.292 ± 1.187 | 0.277 |
| | | Infection probability | 0.417 ± 1.221 | 0.733 |
| Musubi |  |  |  |  |
| | Abdominal pain | (Intercept) | 0.392 ± 0.916 | 0.669 |
| | | Infection probability | 1.872 ± 1.031 | 0.069 |
| | Diarrhoea | (Intercept) | -17.65 ± 1777.31 | 0.992 |
| | | Infection probability | 18.15 ± 1777.31 | 0.992 |
| | Headache | (Intercept) | -0.398 ± 0.916 | 0.664 |
| | | Infection probability | 0.024 ± 0.959 | 0.980 |

<sup>#</sup> Significance codes: 0 – 0.001 (\*\*\*); 0.001 – 0.01 (\*\*); 0.01 – 0.05 (\*); > 0.05 (non-significance)

### Hypotheses 4: Association between infection intensity and side effects

Blood-in-stool, itching/rash, nausea, and pain-when-urinating were not reported as side-effects after treatment, irrespective of infection intensity, in either location (Figure 1, blue). In Bugoto LV abdominal pain, was largely unrelated to infection intensity but a significant negative relationship was likely driven by a skew in the data due to so few people in the higher egg count categories (Figure 2, Table 5). The relationship with diarrhoea, headache, and vomiting was significant but non-linear and peaked within the 121–480 epg range (equivalent to WHO moderate to heavy infection intensities (3)), before declining to zero at higher intensities (Figure 2, Table 5). In Musubi CoG, there was a negligiable relationship observed between infection intensity and headache but a significant negative relationship in the analysis was likely driven by the data skew. A significant non-linear relationship was seen for abdominal pain, diarrhoea, and vomiting, where the prevalence peaked in the 481–600 epg range (equivalent to WHO heavy infection intensities (3)) before also decreasing to zero at the highest intensities (Figure 2, Table 5).

**Table 5.**
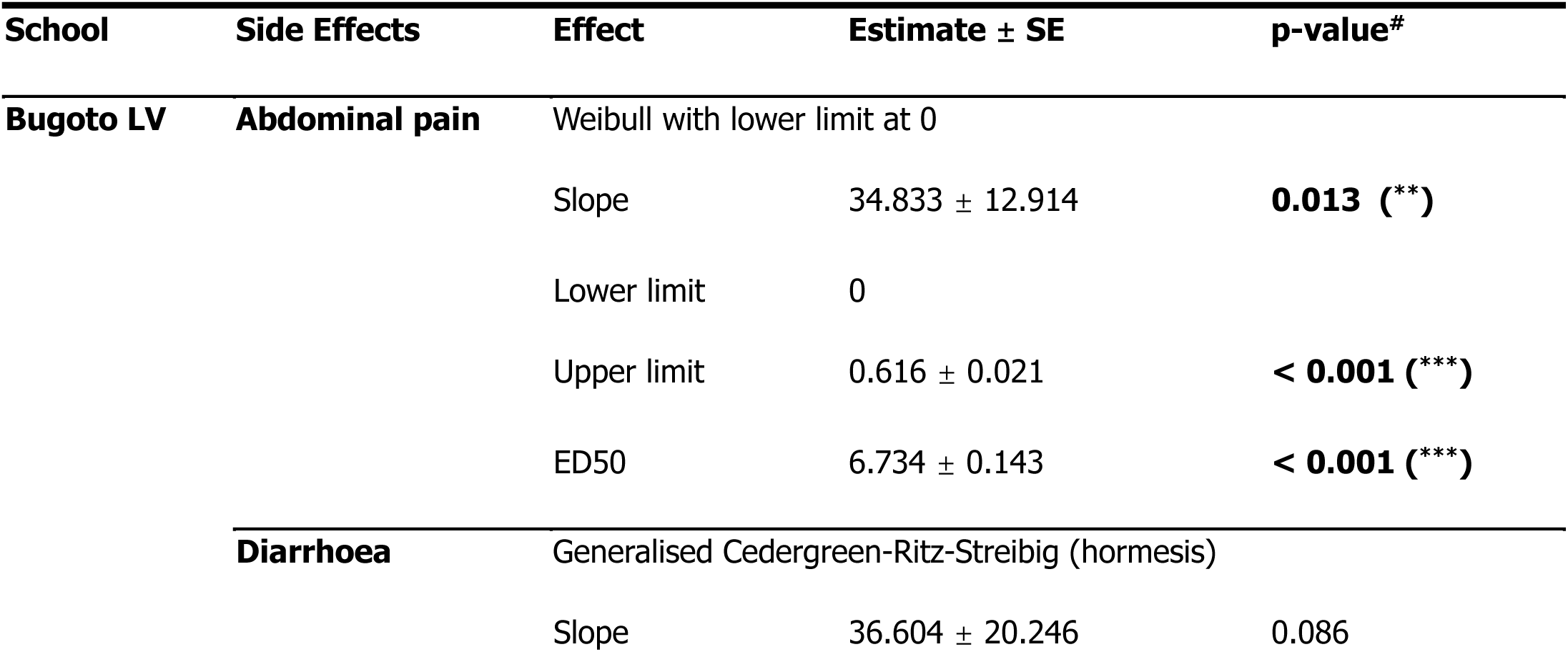
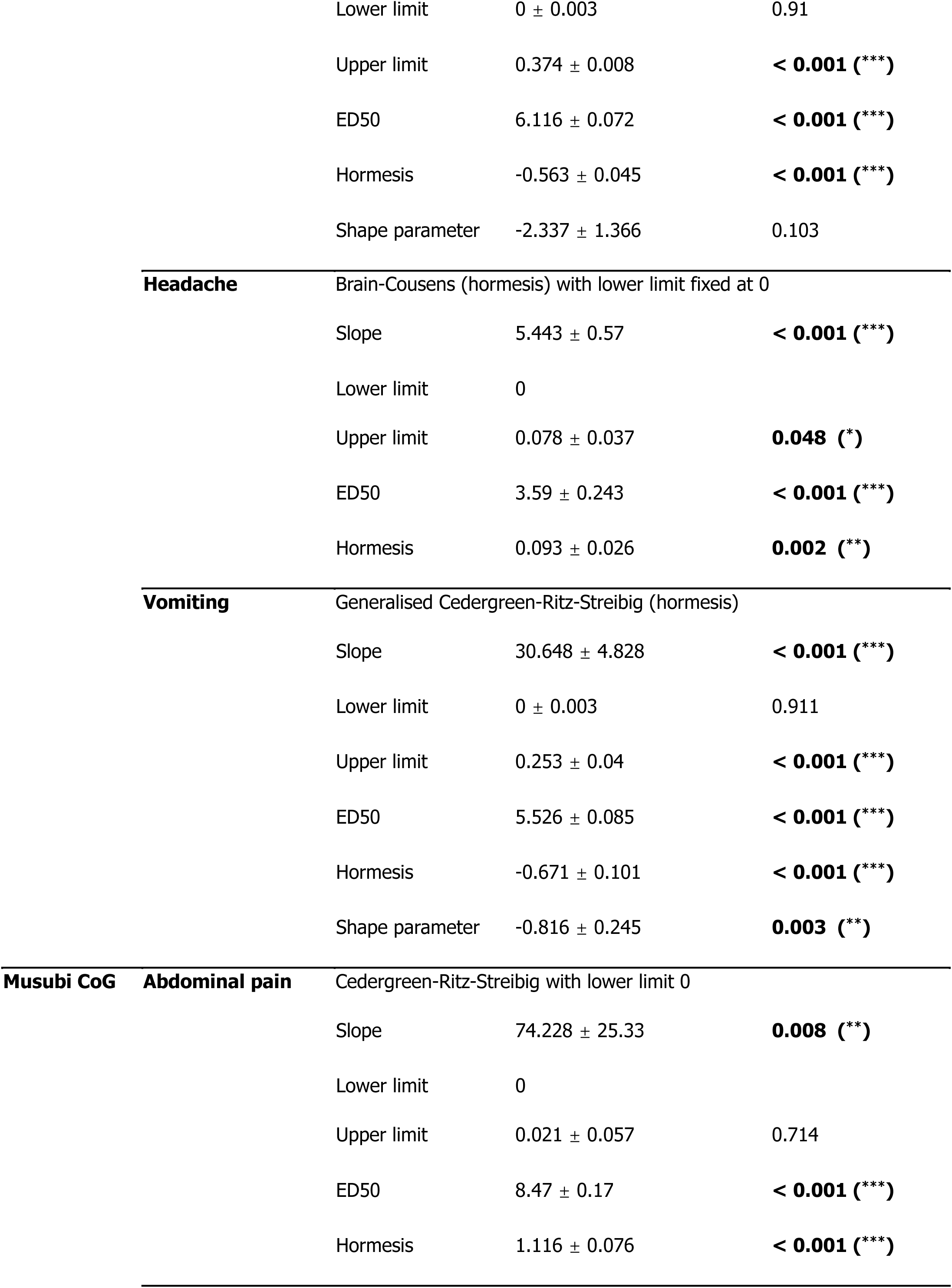
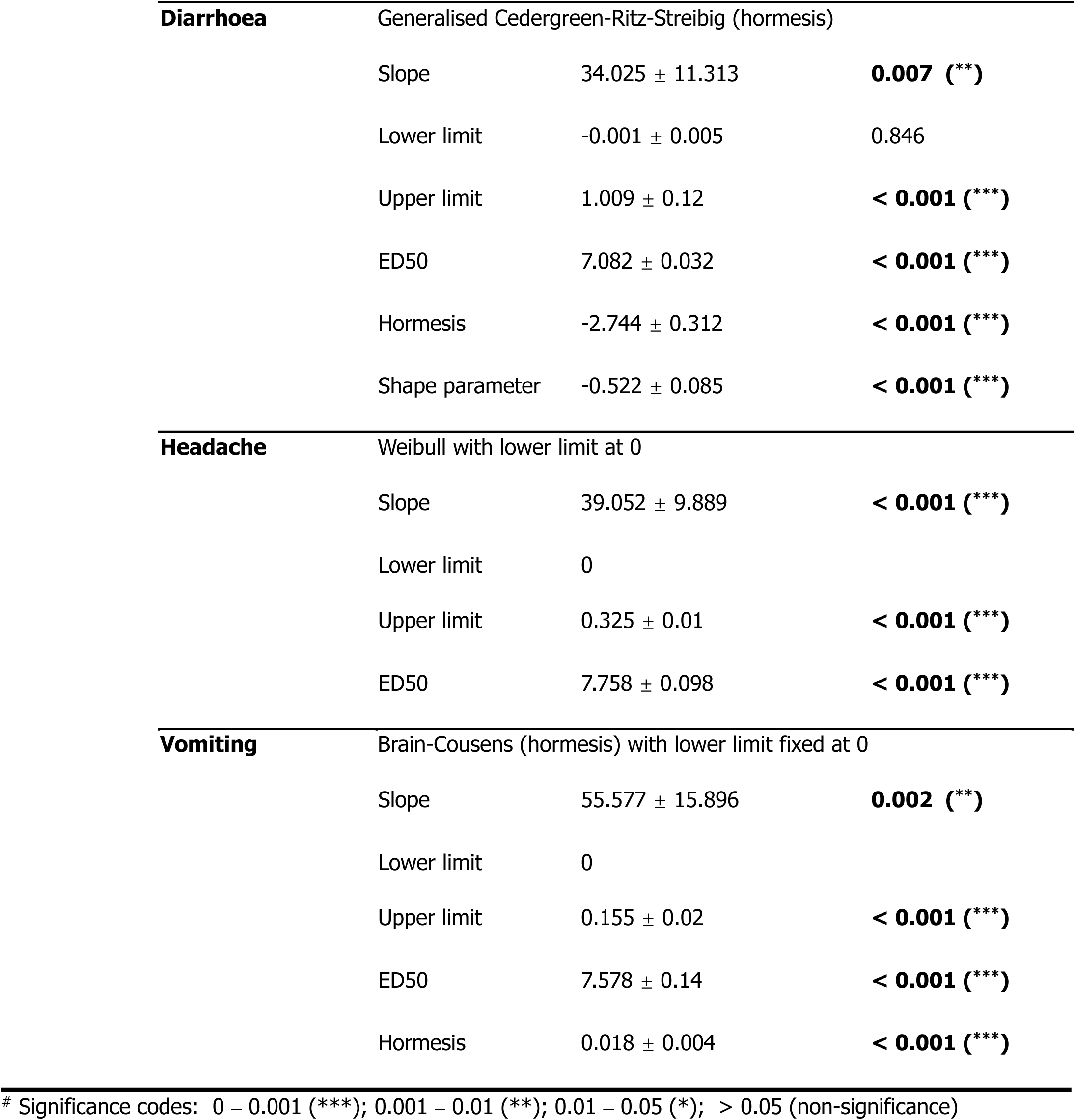
*Schistosoma mansoni* infection intensity dose response results (from the latent class analysis) for side effects where dose response curves could be fitted. The intercept was the group of participants who did not reporting a side effect in every side effect in Bugoto Lake View (LV) and Musubi Church of God (CoG) primary schools in 2004. Statistical significance is indicated as follows: *** p < 0.001, ** p < 0.01, * p < 0.05, and p ≥ 0.05 is considered non-significant.

### Hypotheses 5: Association between side effects and the probability of clearance

Clearance after treatment varied between the two schools (Figure 4) with the mean clearance probability estimated at 82.94% in Bugoto LV and 60.86% in Musubi. The Mann-Whitney U test (W = 2967, *p* = 0.424) indicated that this difference was not statistically significant, suggesting similar distributions in clearance rates between the two schools. At both schools, at the individual-level there was no association between whether an individual successfully cleared infection and whether they reported abdominal pain, diarrhoea, headaches, or vomiting as side effects (Table 6, Figure 4).

**Figure 4.**
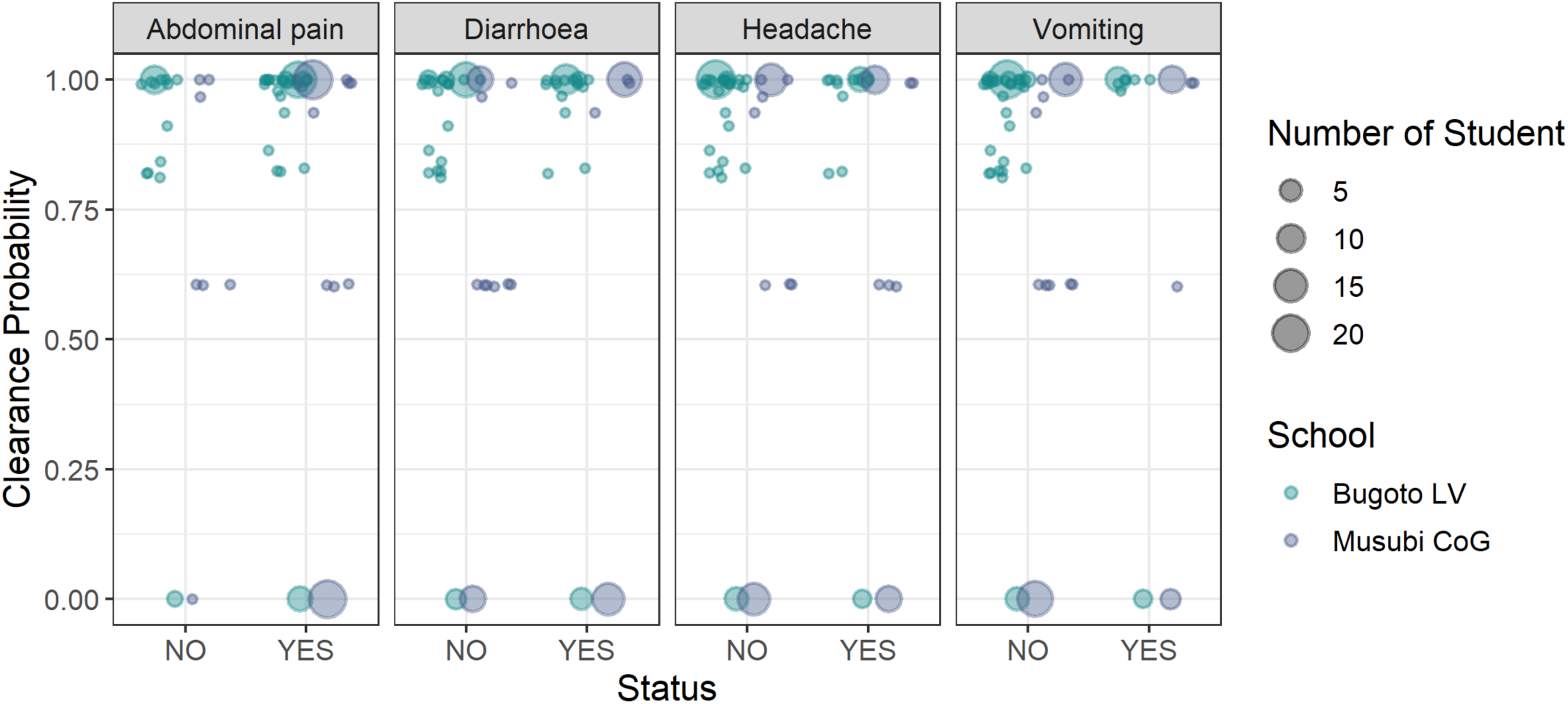
The clearance probability from the latent class analysis compared with not reporting (left – NO) or reporting (right – YES) abdominal pain, diarrhoea, headache, or vomiting as side-effects after praziquantel treatment in Bugoto Lake View (LV) (dark blue) and Musubi Church of God (CoG) (green) primary schools, in 2004. The size of dots relates to the number of participants.

**Table 6.**
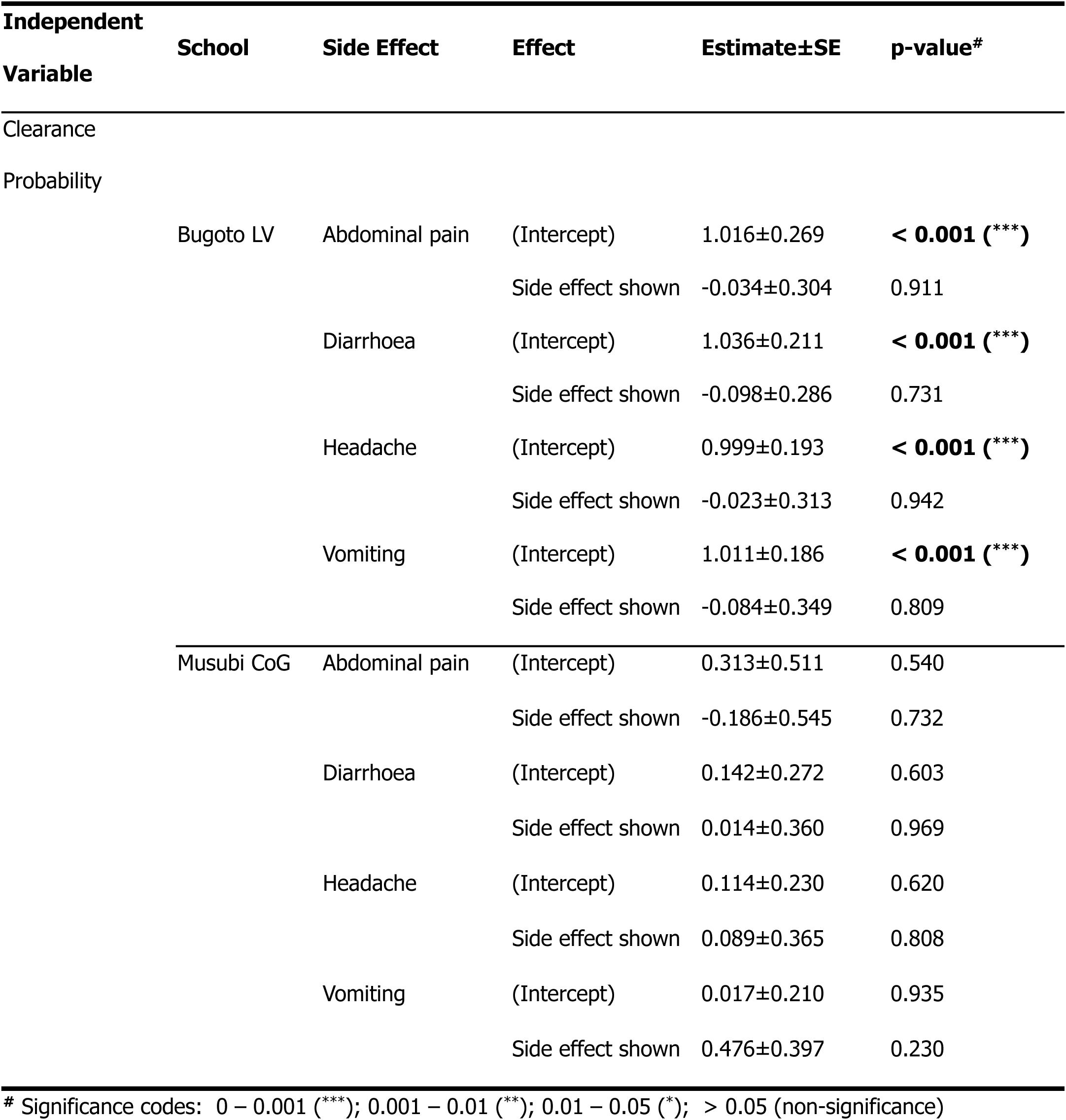
Regression results of the beta distribution models of other side effects (abdominal pain, diarrhoea, headache, and vomiting) and clearance probability after praziquantel treatment with the side effect status as independent variable. The intercept was the group of participants who did not report the side effect in school of Bugoto Lake View (LV) and Musubi Church of God (CoG) primary schools respectively in 2004. Statistical significance is indicated as follows: *** p < 0.001, ** p < 0.01, * p < 0.05, and p ≥ 0.05 is considered non-significant.

| Independent Variable | School | Side Effect | Effect | Estimate±SE | p-value<sup>#</sup> |
| --- | --- | --- | --- | --- | --- |
| Clearance |  |  |  |  |  |
| Probability |  |  |  |  |  |
|  | Bugoto LV | Abdominal pain | (Intercept) | 1.016±0.269 | < **0.001** (***) |
|  |  |  | Side effect shown | -0.034±0.304 | 0.911 |
|  |  | Diarrhoea | (Intercept) | 1.036±0.211 | < **0.001** (***) |
|  |  |  | Side effect shown | -0.098±0.286 | 0.731 |
|  |  | Headache | (Intercept) | 0.999±0.193 | < **0.001** (***) |
|  |  |  | Side effect shown | -0.023±0.313 | 0.942 |
|  |  | Vomiting | (Intercept) | 1.011±0.186 | < **0.001** (***) |
|  |  |  | Side effect shown | -0.084±0.349 | 0.809 |
|  | Musubi CoG | Abdominal pain | (Intercept) | 0.313±0.511 | 0.540 |
|  |  |  | Side effect shown | -0.186±0.545 | 0.732 |
|  |  | Diarrhoea | (Intercept) | 0.142±0.272 | 0.603 |
|  |  |  | Side effect shown | 0.014±0.360 | 0.969 |
|  |  | Headache | (Intercept) | 0.114±0.230 | 0.620 |
|  |  |  | Side effect shown | 0.089±0.365 | 0.808 |
|  |  | Vomiting | (Intercept) | 0.017±0.210 | 0.935 |
|  |  |  | Side effect shown | 0.476±0.397 | 0.230 |

<sup>#</sup> Significance codes: 0 – 0.001 (\*\*\*) ; 0.001 – 0.01 (\*\*); 0.01 – 0.05 (\*); > 0.05 (non-significance)

## Discussion

Praziquantel is administered through MDA to millions of people every year for the control of schistosomaisis morbidity and transmission, aiming to reach the WHO’s EPHP 2030 targets. Despite the extent of treatment, coverage remains low in many places, and side effects are reported as reasons for low uptake (38). To achieve EPHP, MDA has largely been scaled up to include a wider proportion of the community than just school-aged children (18). MDA remains a major part of the WHO’s ambitious goals of EPHP by 2030, and an improved evidence base is required to enable improved sensitisation and accurate information to be disseminated to communities endemic for schistosomaisis. In this study we tested the relationship between *S. mansoni* infection prevalence, intensity, and pre-treatment self reported symptoms and post-treatment self reported side effects. We also tested whether side effects were related to drug efficacy four weeks after praziquantel MDA. These data were collected in the very early years of Uganda’s MDA programme which has now been running for over 20 years. Our analyses do not entirely support published relationships (37,53) that suggest severity of praziquantel treatment side effects are positively correlated with infection intensity of *S. mansoni.* Instead, we show that this relationship is variable across settings and often non-linear, often peaking in prevalence at sub-maximum egg counts. Our findings support the general concensus that morbidity is rarely linearly associated with infection intensity. Our results also do not support the hypothesis that side effects are related to subsequent parasite clearance probability. Notably, despite Musubi CoG being drug-naïve prior to treatment and Bugoto LV having received one prior round of MDA, praziquantel efficacy was comparable between the two schools. Our results indicate that interactions between infection status, symptoms, side effects and treatment efficacy are complex making it difficult to predict individual responses to praziquantel treatment with certainty. Below we discuss each of our hypothese in more detail, highlight the implications of our findings, and detail the limitations of this study and further research that may help understand any possible relationships between *S. mansoni* and praziquantel side effects.

### Hypotheses 1 & 2

Our analyses tested whether the probability of infection was associated with the reporting of symptoms (hypothesis 1), and whether infection intensity before treatment had an effect on symptom reporting (hypothesis 2). However, our ability to test hypothesis 1 may have been limited, particularly in Musubi, by the strong imbalance in the data, as over 90% of participants were classified as infected pre-treatment, reducing the statistical power to detect an association. While we did not find support for hypothesis 1, this result should be interpreted with caution given the highly skewed infection status distribution. For hypothesis 2, the relationship between symptoms and infection intensity was complex, varied between schools, and was often non-linear. Reports of blood-in-stool did increase with increasing infection intensity in both schools up to a point, after which there was a strong decline in the association. This finding suggests that blood in stool may be more strongly linked to *S. mansoni* infection than other conditions, possibly due to the parasite eggs penetrating and damaging the gut lining to be excreted (4,10,15). It may be that at the highest infection intensities, egg excretion may actually be more efficient and may indicate less morbidity per egg, and so less blood per egg. There is increasing recognition that infection intensity as measured by eggs excreted in stool is not linearly related to morbidity (41,54). In the cases of nausea and pain-when-urinating at Bugoto LV, and abdominal pain, diarrhoea, nausea and itching/rash at Musubi CoG, prevalence of these symptoms was notable even in those who were classified as not infected (i.e. 0 epg and with negative or low antigen scores and therefore more likely to be estimated as uninfected as by the model). This suggests that these are not only associated with schistosomiasis infection – and are generally not related to infection intensity, and may indicate that whatever is causing these morbidities is a stronger driver or is at least not exacerbated by schistosomiasis. This lack of association may reflect limitations in how infection intensity is measured. Even though we used a latent class analysis model that incorporates both egg and antigen data to infer infection status, the model still relies on egg shedding as a proxy for worm burden. However, egg output can vary widely between individuals with similar worm loads. Other approaches, such as sibship reconstruction (55), may help provide a more direct estimate of worm burden by analysing genetic relationships between excreted parasite eggs. Although these methods are also constrained by the need for sufficient egg samples, they could offer more accurate insights into the true underlying infection dynamics.

The decline in symptom prevalence at higher egg counts should be interpreted with caution. In particular, the small number of individuals in the highest infection categories may have exaggerated apparent associations, especially for symptoms that were infrequently reported. This issue recurred across multiple symptoms, suggesting that caution is needed when interpreting non-linear trends. For example, in the case of pain when urinating, the decline observed at high infection intensities may reflect random variation amplified by the low number of cases and the overall low reporting frequency of this symptom (∼25%).

Symptoms of infection with soil transmitted helminths can also resemble *S. mansoni* infection (4,12–15,56), and they are known to often co-occur in the same communities and individuals (44,57). The possibility of co-infection with *S. mansoni* and STH might explain some of the symptoms reported prior to treatment where no relationship was found with infection or intensity. Bugoto LV also received Albendazole treatment in 2003 when they received their initial MDA, therefore it is possible that STH – *S. mansoni* co-infection was less prevalent, potentially explaining why participants from Bugoto LV reported fewer pre-treatment symptoms. Additionally, regions with a high incidence of schistosomiasis often overlap with areas endemic for malaria, suggesting that malaria should also be considered as a potential contributing factor of symptom severity (58–60), likely impacting the presence and/or reporting of symptoms. Additionally, reports indicate that villages surrounding Lake Victoria frequently experience a high burden of diarrhoeal diseases (59), largely due to inadequate sanitary infrastructure and contamination of water sources. High levels of faecal contamination in the soil and in lake water, particularly during the wet season, contribute to the spread of these diseases. Limited access to improved sanitation facilities and reliance on untreated lake water further exacerbate the problem, increasing the risk of infections among local communities. Symptoms of infections such as soil-transmitted helminths (STH), malaria, diarrhoeal diseases, etc., can be similar to *S. mansoni* infection, albeit commonly much more acute. While these co-infections may explain the lack of relationships found in some cases, our results highlight that specific symptoms, such as blood in stool and nausea, were significantly associated with *S. mansoni* infection intensity, underscoring the importance of considering multiple factors when interpreting symptom patterns.

### Hypotheses 3 & 4

Our analyses next tested whether there was a relationship between the probability of infection and the reporting of side effects (hypothesis 3), or infection intensity and the reporting of side effects (hypothesis 4). We found different outcomes for each school. More participants from Musubi reported side effects than those at Bugoto LV. At Bugoto LV, diarrhoae, headache and vomiting increased initially with increasing infection prevalence. Adominal pain did mildly increase in prevalence with at moderate infection intensties but was also prevalent in 50% of uninfected children. The significant negative association observed between abdominal pain and infection intensity may also reflect the effects of data skew. Given the low number of individuals in the highest egg count categories, and the high background prevalence of abdominal pain even among uninfected individuals, statistical significance in this case is likely sensitive to small fluctuations in reporting. This again highlights the difficulty in drawing strong conclusions from symptom data in strata with limited sample size. Children at Musubi CoG reported increasing amounts of diarrhoae and vomiting with increasing infection intensities. Headaches were likely also associated with other causes. The differences of pre-treatment infection intensity and post-treatment side effect reports between the two schools could be attributed to the fact that Musubi CoG was praziquantel-naïve in 2004 when these data were collected, while Bugoto LV had already participated in one praziquantel MDA in 2003 (61). If praziquantel is effective at treating *S. mansoni* infections, and the side effects are a function of worms dying as has been previously suggested, then this may explain our observed differences in side effect reporting. However, since our analysis accounts for infection intensity by comparing participants within the same intensity bins, the overall lower infection intensities in Bugoto LV should not, in itself, drive these differences. If the number of dying worms were the primary driver of side effects, we would expect to see a consistent association between heavier infections and increased side effects across both schools. Despite some students from Bugoto LV still harbouring heavy infections one year after MDA started, we did not observe this pattern (28,43). This suggests that additional factors such as prior drug exposure, host immune responses, or co-infections may also play a role in shaping the differences in reported side effects.

Our surveys recorded symptoms and side effects in a binary manner, which limits the ability to capture detailed variations, and it may be that the severity of specific symptoms does increase with higher infection intensities. To improve upon this, future research could employ scales to provide a more nuanced understanding of symptom and side effect severity and duration, as well as their impact on health-related quality of life (HRQoL). A recent study highlighted that integrating HRQoL assessments offers valuable insights into the broader effects of treatment on patients’ well-being, but demonstrated a complex relationship between infectons, symptons and HRQoL (45). It is also possible that participants who experienced severe symptoms pre-treatment may be less likely to recognise, classify and/or report side-effects after treatment, as they already experienced them at the time of treatment and they are not ‘new’ drug induced side effects. Again, a scale of the severity of symptoms and side effects may help to elucidate if pre-treatment symptoms affect post-treatment side effect reporting. We had hoped to address this question, but upon further investigation, our data were not appropriate to statistically analyse this and a more systematic method of recording pre treatment symptoms and post-treatment side effect data is needed.

Without these improved data it is not possible to delineate whether a participant had not reported the side effect because it was not there, or because it was not worse than the symptom they may have been experiencing prior to treatment.

### Hypothesis 5

We tested whether the occurrence of side effects was related to the probability of infection clearance after praziquantel treatment. Our analysis demonstrated a high clearance rate among participants from both Bugoto LV and Musubi CoG schools following treatment. However, there was no significant relationship between the reporting of abdominal pain, diarrhoea, or headache as side effects and successful clearance (and the other side-effects could not be fitted to the models). This indicates that the presence of these specific side effects does not predict the efficacy of praziquantel treatment in clearing *S. mansoni* infections in these participants and therefore that side effects experienced post-treatment can not be used as reliable indicators of successful worm clearance. This is in contrast to previous studies which suggested that severe side effects might be indicative of substantial worm death, leading to higher treatment efficacy (21,34,40).

#### Limitations

A key limitation in our analysis was the small number of participants with very high pre-treatment infection intensities, which is epidemiologically resonable but resulted in sparse data within higher intensity categories. This led to a strong skew towards zero in the proportion of individuals reporting symptoms or side effects across infection intensity categories, particularly at the upper end of the scale. In the figure associated with Hypotheses 2 and 4 (figure 2), most symptom and side effect reporting curves decline rapidly to zero at higher infection intensities, potentially reflecting data sparsity rather than true biological patterns. While this does not necessarily indicate errors in the data, it highlights potential complexities in infection dynamics, symptom reporting, or measurement variability that may require alternative analytical approaches in future studies.

We also observed that some participants experienced vomiting after taking the medication, a concern that was also documented at the start of the Schistosomiasis Control Initiative (SCI), but our current survey did not track whether they continued to experience this side effect after subsequent doses. Future research should investigate whether vomiting and its timing impact treatment outcomes by including follow-up assessments after drug re-administration.

Participants who remained *S. mansoni* positive one week after treatment were retreated with an additional praziquantel dose. While this is not ideal for the questions we are asking from the data here, it occurred to check when eggs were reducing post treatment and when drug efficacy measures should be made. It demonstrated that a significant proportion of treated individuals were still egg positive one week after treatment and is now well known drug efficacy measurements need to occur longer after treatment. As so many children were still infected with >100 epg, many were therefore retreated which will likely have strongly influenced our drug efficacy measurements at four weeks, as many participants received two doses rather than one. Although this does not impact the association between infection intensity and side effects (which were reported after the first dose), it may have obscured potential relationships between infection intensity and drug efficacy, contributing to higher overall clearance rates observed than would have occurred with just a single treatment. Future studies that follow current treatment routines of a single treatment will be essential for disentangling these effects and better understanding the relationship between infection intensity, side effects, and drug efficacy.

#### Further work

In endemic areas where rapid reinfection is common (62,63), repeated MDA remains necessary. Understanding the factors influencing side effects and reinfection dynamics is crucial for optimizing treatment strategies. Longitudinal studies tracking the same individuals over time could provide valuable insights into whether side effects diminish with repeated treatments and whether treatment history affects symptom severity. These findings could support evidence-based health education campaigns, helping individuals and communities make informed decisions about MDA participation while ensuring their concerns and well-being are appropriately addressed.

### Summary and Conclusions

Although praziquantel is effective against *S. mansoni*, it can cause temporary side effects such as abdominal pain, diarrhoae, and vomiting. While these symptoms are generally short-lived, they can be distressing for individuals undergoing treatment. Ensuring that individuals are informed about potential side effects and their transient nature is essential for ethical reasons as well as potentially improving treatment adherence by preparing people accordingly. There is not evidence that side-effects are positively associated with infection intensites and so using this explantion to improve return treatment would not be valid. However, conversely, clarifying that these ailments do not indicate infection status may improve uptake by removing stigma but further studies on such downstream impacts are required. Simple interventions such as co-administration of medications like paracetamol for headache and abdominal pain, or antihistamines for itching and rash, could help mitigate discomfort.

Our study investigated the relationship between pre-treatment infection intensity and the presence of certain side effects, which might serve as indirect evidence of worm death following praziquantel treatment. Participants from Bugoto LV, where a previous round of treatment had been administered, reported fewer side effects than those from Musubi CoG, where treatment was being delivered for the first time. Further research will help understand if such differences are due to different previous praziquantel exposure or other factors and how treatment history may influence the frequency or severity of reported side effects, especially now as control programmes have now been running for many years/decades, which may result in different findings in comparison to this study which analyses data from the start of Uganda’s control programme.

Mass drug administration (MDA) remains the cornerstone of schistosomiasis control and plays a key role in achieving the WHO 2030 targets for eliminating the disease as a public health problem. However, despite extensive treatment efforts, challenges remain, particularly in addressing concerns about side effects, treatment adherence, and reinfection dynamics. As control programmes progress, managing side effects and improving community engagement may be critical to sustaining participation in MDA initiatives.

## Data Availability

All data and anonymised code are available at https://github.com/iamjessclark/symptomsSideEffects.git

## Acknowledgements

We are sincerely grateful to the National and District officials, and drivers from the Vector Control Division of the Ministry of Health for their support. We also thank head teachers and teachers from Bugoto LV and Musubi CoG for their assistance in the field. Our appreciations go to the village health teams and village local council chairpersons for their efforts in mobilizing the community to participate in this study. Lastly, we are deeply grateful to the participants who provided samples and data for this research.

## References

1. Vos T, Flaxman AD, Naghavi M, Lozano R, Michaud C, Ezzati M, et al. Years lived with disability (YLDs) for 1160 sequelae of 289 diseases and injuries 1990–2010: a systematic analysis for the Global Burden of Disease Study 2010. The Lancet. 2012;380(9859): 2163–2196. 10.1016/S0140-6736(12)61729-2.

2. Chitsulo L, Loverde P, Engels D. Focus: Schistosomiasis. Nature Reviews Microbiology 2004 2:1. 2004;2(1): 12–12. 10.1038/nrmicro801.

3. World Health Organization. WHO GUIDELINE on control and elimination of human schistosomiasis. 2022.

4. Da Silva L, Chieffi P, Carrilho F. Schistosomiasis mansoni - Clinical features. Gastroenterología y Hepatología. 2005;28(1): 30–39. 10.1157/13070382.

5. Enk M, Lima A, Drummond S, Schall V, Coelho P. The effect of the number of stool samples on the observed prevalence and the infection intensity with *Schistosoma mansoni* among a population in an area of low transmission. Acta tropica. 2008;108(2–3): 222–228. 10.1016/J.ACTATROPICA.2008.09.016.

6. Colley DG, Binder S, Campbell C, King CH, Tchuenté LAT, N’Goran EK, et al. A Five-Country Evaluation of a Point-of-Care Circulating Cathodic Antigen Urine Assay for the Prevalence of *Schistosoma mansoni*. The American Journal of Tropical Medicine and Hygiene. 2013;88(3): 426. 10.4269/AJTMH.12-0639.

7. Clark J, Moses A, Nankasi A, Faust CL, Adriko M, Ajambo D, et al. Translating From Egg- to Antigen-Based Indicators for *Schistosoma mansoni* Elimination Targets: A Bayesian Latent Class Analysis Study. Frontiers in tropical diseases. 2022;3. 10.3389/FITD.2022.825721.

8. Clements MN, Corstjens PLAM, Binder S, Campbell CH, De Dood CJ, Fenwick A, et al. Latent class analysis to evaluate performance of point-of-care CCA for low-intensity *Schistosoma mansoni* infections in Burundi. Parasites & Vectors. 2018;11(1). 10.1186/S13071-018-2700-4.

9. Prada JM, Touloupou P, Adriko M, Tukahebwa EM, Lamberton PHL, Hollingsworth TD. Understanding the relationship between egg- and antigen-based diagnostics of *Schistosoma mansoni* infection pre- and post-treatment in Uganda. Parasites and Vectors. 2018;11(1): 1–8. 10.1186/S13071-017-2580-Z/FIGURES/4.

10. Costain AH, MacDonald AS, Smits HH. Schistosome Egg Migration: Mechanisms, Pathogenesis and Host Immune Responses. Frontiers in Immunology. 2018;9: 3042. 10.3389/FIMMU.2018.03042.

11. Doherty JF, Moody AH, Wright SG. Lesson of the Week: Katayama fever: an acute manifestation of schistosomiasis. BMJ. 1996;313(7064): 1071–1072. 10.1136/BMJ.313.7064.1071.

12. Peterson WP, von Lichtenberg F. Studies on Granuloma Formation. IV. The Journal of Immunology. 1965;95(5).

13. Midzi N, Sangweme D, Zinyowera S, Mapingure MP, Brouwer KC, Kumar N, et al. Efficacy and side effects of praziquantel treatment against *Schistosoma haematobium* infection among primary school children in Zimbabwe. Transactions of the Royal Society of Tropical Medicine and Hygiene. 2008;102(8): 759–766. 10.1016/J.TRSTMH.2008.03.010/2/102-8-759-TBL005.GIF.

14. Cao J, Liu WJ, Xu XY, Zou XP. Endoscopic findings and clinicopathologic characteristics of colonic schistosomiasis: A report of 46 cases. World Journal of Gastroenterology : WJG. 2010;16(6): 723. 10.3748/WJG.V16.I6.723.

15. Bharti AR, Weidner N, Ramamoorthy S. Chronic Schistosomiasis in a Patient with Rectal Cancer. The American Journal of Tropical Medicine and Hygiene. 2009;80(1): 1–2. 10.4269/AJTMH.2009.80.1.

16. Verjee MA. Schistosomiasis: Still a Cause of Significant Morbidity and Mortality. Research and Reports in Tropical Medicine. 2019;10: 153. 10.2147/RRTM.S204345.

17. Andrews P, Thomas H, Pohlke R, Seubert Jr. Praziquantel. Medicinal Research Reviews. 1983;3(2): 147–200. 10.1002/MED.2610030204.

18. WHO. Ending the neglect to attain the Sustainable Development Goals: a road map for neglected tropical diseases 2021–2030. 2020.

19. Colley DG, Bustinduy AL, Secor WE, King CH. Human schistosomiasis. The Lancet. 2014;383(9936): 2253–2264. 10.1016/S0140-6736(13)61949-2.

20. WHO. Ending the neglect to attain the Sustainable Development Goals: a road map for neglected tropical diseases 2021–2030. Geneva: World Health Organization (https://www.who.int/neglected_diseases/Revised-DraftNTD-Roadmap-23Apr2020.pdf). 2020.

21. Cioli D, Pica-Mattoccia L. Praziquantel. Parasitology Research. 2003;90: S3–S9.

22. Doenhoff MJ, Cioli D, Utzinger J. Praziquantel: Mechanisms of action, resistance and new derivatives for schistosomiasis. Current Opinion in Infectious Diseases. 2008;21(6): 659–667. 10.1097/QCO.0B013E328318978F.

23. Wang W, Wang L, Liang YS. Susceptibility or resistance of praziquantel in human schistosomiasis: a review. Parasitology Research 2012 111:5. 2012;111(5): 1871–1877. 10.1007/S00436-012-3151-Z.

24. Vale N, Gouveia MJ, Rinaldi G, Brindley PJ, Gärtner F, Da Costa JMC. Praziquantel for schistosomiasis: Single-drug metabolism revisited, mode of action, and resistance. Antimicrobial Agents and Chemotherapy. 2017;61(5). 10.1128/AAC.02582-16.

25. Seto EYW, Wong BK, Lu D, Zhong B. Human Schistosomiasis Resistance to Praziquantel in China: Should We Be Worried? The American Journal of Tropical Medicine and Hygiene. 2011;85(1): 74. 10.4269/AJTMH.2011.10-0542.

26. Fukushige M, Chase-Topping M, Woolhouse MEJ, Mutapi F. Efficacy of praziquantel has been maintained over four decades (from 1977 to 2018): A systematic review and meta-analysis of factors influence its efficacy. PLOS Neglected Tropical Diseases. 2021;15(3): e0009189. 10.1371/JOURNAL.PNTD.0009189.

27. Crellen T, Walker M, Lamberton PHL, Kabatereine NB, Tukahebwa EM, Cotton JA, et al. Reduced Efficacy of Praziquantel Against *Schistosoma mansoni* Is Associated With Multiple Rounds of Mass Drug Administration. Clinical Infectious Diseases. 2016;63(9): 1151–1159. 10.1093/CID/CIW506.

28. Clark J, Moses A, Nankasi A, Faust CL, Moses A, Ajambo D, et al. Reconciling Egg- and Antigen-Based Estimates of *Schistosoma mansoni* Clearance and Reinfection: A Modeling Study. Clinical Infectious Diseases. 2021; 10.1093/CID/CIAB679.

29. MT I, RM O, TN C, DU O, AG R. Prevention and control of schistosomiasis: a current perspective. Research and reports in tropical medicine. 2014;2014(5): 65. 10.2147/RRTM.S44274.

30. Gönnert R, Andrews P. Praziquantel, a new broad-spectrum antischistosomal agent. Zeitschrift für Parasitenkunde 1977 52:2. 1977;52(2): 129–150. 10.1007/BF00389899.

31. Greenberg RM. Are Ca2+ channels targets of praziquantel action? International Journal for Parasitology. 2005;35(1): 1–9. 10.1016/J.IJPARA.2004.09.004.

32. Chulkov EG, Rohr CM, Marchant JS. Praziquantel activates a native cation current in *Schistosoma mansoni*. Frontiers in Parasitology. 2023;2: 1285177. 10.3389/FPARA.2023.1285177.

33. Trienekens SCM, Faust CL, Meginnis K, Pickering L, Ericssonid O, Nankasi A, et al. Impacts of host gender on *Schistosoma mansoni* risk in rural Uganda—A mixed-methods approach. PLOS Neglected Tropical Diseases. 2020;14(5): e0008266. 10.1371/JOURNAL.PNTD.0008266.

34. Park SK, Friedrich L, Yahya NA, Rohr C, Chulkov EG, Maillard D, et al. Mechanism of praziquantel action at a parasitic flatworm ion channel. bioRxiv. 2021; 2021.03.09.434291. 10.1101/2021.03.09.434291.

35. Jaoko WG, Muchemi G, Oguya FO. Praziquantel side effects during treatment of *Schistosoma mansoni* infected pupils in Kibwezi, Kenya - PubMed. East Afr Med J. 1996;73(8): 499–501.

36. Hoekstra PT, Casacuberta-Partal M, Lieshout L van, Corstjens PLAM, Tsonaka R, Assaré RK, et al. Efficacy of single versus four repeated doses of praziquantel against *Schistosoma mansoni* infection in school-aged children from Côte d’Ivoire based on Kato-Katz and POC-CCA: An open-label, randomised controlled trial (RePST). PLOS Neglected Tropical Diseases. 2020;14(3): e0008189. 10.1371/JOURNAL.PNTD.0008189.

37. Berhe N, Gundersen S, Abebe F, Birrie H, Medhin G, Gemetchu T. Praziquantel side effects and efficacy related to *Schistosoma mansoni* egg loads and morbidity in primary school children in north-east Ethiopia. Acta tropica. 1999;72(1): 53–63. 10.1016/S0001-706X(98)00084-9.

38. Adriko M, Faust CL, Carruthers L V., Moses A, Tukahebwa EM, Lamberton PHL. Low Praziquantel Treatment Coverage for *Schistosoma mansoni* in Mayuge District, Uganda, Due to the Absence of Treatment Opportunities, Rather Than Systematic Non-Compliance. Tropical Medicine and Infectious Disease. 2018;3(4). 10.3390/TROPICALMED3040111.

39. Ssali A, Pickering L, Nalwadda E, Mujumbusi L, Seeley J, Lamberton PHL. Schistosomiasis messaging in endemic communities: Lessons and implications for interventions from rural Uganda, a rapid ethnographic assessment study. PLOS Neglected Tropical Diseases. 2021;15(10): e0009893. 10.1371/JOURNAL.PNTD.0009893.

40. Kabatereine NB, Kemijumbi J, Ouma JH, Sturrock RF, Butterworth AE, Madsen H, et al. Efficacy and side effects of praziquantel treatment in a highly endemic *Schistosoma mansoni* focus at Lake Albert, Uganda. Transactions of The Royal Society of Tropical Medicine and Hygiene. 2003;97(5): 599–603. 10.1016/S0035-9203(03)80044-5.

41. Wiegand RE, Secor WE, Fleming FM, French MD, King CH, Montgomery SP, et al. Control and Elimination of Schistosomiasis as a Public Health Problem: Thresholds Fail to Differentiate Schistosomiasis Morbidity Prevalence in Children. Open Forum Infectious Diseases. 2021;8(7). 10.1093/ofid/ofab179.

42. Lamberton PHL, Hogan SC, Kabatereine NB, Fenwick A, Webster JP. In vitro praziquantel test capable of detecting reduced in vivo efficacy in *Schistosoma mansoni* human infections. American Journal of Tropical Medicine and Hygiene. 2010;83(6): 1340–1347. 10.4269/ajtmh.2010.10-0413.

43. Lamberton PHL, Kabatereine NB, Oguttu DW, Fenwick A, Webster JP. Sensitivity and Specificity of Multiple Kato-Katz Thick Smears and a Circulating Cathodic Antigen Test for *Schistosoma mansoni* Diagnosis Pre- and Post-repeated-Praziquantel Treatment. PLoS Neglected Tropical Diseases. 2014;8(9). 10.1371/JOURNAL.PNTD.0003139.

44. Kabatereine N, Brooker S, Koukounari A, Kazibwe F, Tukahebwa E, Fleming F, et al. Impact of a national helminth control programme on infection and morbidity in Ugandan schoolchildren. Bulletin of the World Health Organization. 2007;85(2): 91–99. 10.1590/S0042-96862007000200006.

45. Alonso S, Arinaitwe M, Atuhaire A, Nankasi AB, Prada JM, McIntosh E, et al. The short-term impact of *Schistosoma mansoni* infection on health-related quality of life: implications for current elimination policies. Proceedings B. 2024;291(2024). 10.1098/RSPB.2024.0449.

46. R Core Team. R: A Language and Environment for Statistical Computing. 2021.

47. Wickham H, Averick M, Bryan J, Chang W, D’ L, Mcgowan A, et al. Welcome to the Tidyverse. Journal of Open Source Software. 2019;4(43): 1686. 10.21105/JOSS.01686.

48. Hartig F, Lohse L. Residual Diagnostics for Hierarchical (Multi-Level / Mixed) Regression Models. 2021.

49. Denwood MJ. runjags: An R Package Providing Interface Utilities, Model Templates, Parallel Computing Methods and Additional Distributions for MCMC Models in JAGS. Journal of Statistical Software. 2016;71: 1–25. 10.18637/JSS.V071.I09.

50. Brooks ME, Kristensen K, Van Benthem KJ, Magnusson A, Berg CW, Nielsen A, et al. glmmTMB Balances Speed and Flexibility Among Packages for Zero-inflated Generalized Linear Mixed Modeling.

51. Ritz C, Baty F, Streibig JC, Gerhard D. Dose-Response Analysis Using R. PLOS ONE. 2015;10(12): e0146021. 10.1371/JOURNAL.PONE.0146021.

52. Smithson M, Verkuilen J. A better lemon squeezer? Maximum-likelihood regression iith Beta-distributed dependent variables. 2006;11(1): 54–71. 10.1037/1082-989X.11.1.54.

53. Stelma FF, Talla I, Sow S, Kongs A, Niang M, Polman K, et al. Efficacy and side effects of praziquantel in an epidemic focus of *Schistosoma mansoni*. American Journal of Tropical Medicine and Hygiene. 1995;53(2): 167–170. 10.4269/ajtmh.1995.53.167.

54. Wiegand RE, Fleming FM, de Vlas SJ, Odiere MR, Kinung’hi S, King CH, et al. Defining elimination as a public health problem for schistosomiasis control programmes: beyond prevalence of heavy-intensity infections. The Lancet Global Health. 2022;10(9): e1355–e1359. 10.1016/S2214-109X(22)00287-X.

55. Neves MI, Webster JP, Walker M. Estimating helminth burdens using sibship reconstruction. Parasites and Vectors. 2019;12(1): 1–12. 10.1186/S13071-019-3687-1/FIGURES/6.

56. Jourdan PM, Lamberton PHL, Fenwick A, Addiss DG. Soil-transmitted helminth infections. The Lancet. 2018. p. 252–265. 10.1016/S0140-6736(17)31930-X.

57. Kolaczinski JH, Onapa AW, Ndyomugyenyi R, Brooker S. Neglected Tropical Diseases and Their Control in Uganda. Situational analysis and needs assessment report. 2006;(April).

58. Balen J, Stothard JR, Kabatereine NB, Tukahebwa EM, Kazibwe F, Whawell S, et al. Morbidity due to *Schistosoma mansoni*: an epidemiological assessment of distended abdomen syndrome in Ugandan school children with observations before and 1-year after anthelminthic chemotherapy. Transactions of the Royal Society of Tropical Medicine and Hygiene. 2006;100(11): 1039–1048. 10.1016/j.trstmh.2005.12.013.

59. Muyodi FJ, Hecky RE, Kitamirike JM, Odong R. Water quality and health conditions in Lake Victoria region, Uganda. 2005;(December): 178–202.

60. World Health Organization. World malaria report 2023. 2023. https://www.wipo.int/amc/en/mediation/

61. Kabatereine NB, Tukahebwa E, Kazibwe F, Namwangye H, Zaramba S, Brooker S, et al. Progress towards countrywide control of schistosomiasis and soil-transmitted helminthiasis in Uganda. Transactions of the Royal Society of Tropical Medicine and Hygiene. 2006;100(3): 208–215. 10.1016/j.trstmh.2005.03.015.

62. Sircar AD, Mwinzi PNM, Onkanga IO, Wiegand RE, Montgomery SP, Evan Secor W. *Schistosoma mansoni* Mass Drug Administration Regimens and Their Effect on Morbidity among Schoolchildren over a 5-Year Period— Kenya, 2010–2015. The American Journal of Tropical Medicine and Hygiene. 2018;99(2): 362–369. 10.4269/AJTMH.18-0067.

63. Kittur N, Binder S, Campbell CH, King CH, Kinung’Hi S, Olsen A, et al. Defining Persistent Hotspots: Areas That Fail to Decrease Meaningfully in Prevalence after Multiple Years of Mass Drug Administration with Praziquantel for Control of Schistosomiasis. The American Journal of Tropical Medicine and Hygiene. 2017;97(6): 1810. 10.4269/AJTMH.17-0368.

