## supplementaryMaterials for "The associations between *Schistosoma mansoni* infection, pre-treatment symptoms, praziquantel side effects, and treatment efficacy in Ugandan school-aged children"

**Supplementary Materials**

**Table S1.**	The number of students in each school participating in the research in 2004.

|  | Number of tested students | Number of students included in analysis |
| --- | --- | --- |
| Bugoto LV | 123 | 94 |
| Musubi CoG | 68 | 68 |
| Total | 191 | 162 |

In Bugoto LV, 123 children were tested and in Musubi CoG, 68 children were tested. Of the 94 children in Bugoto LV and all of the children in Musubi CoG, their data can be used for HMM.

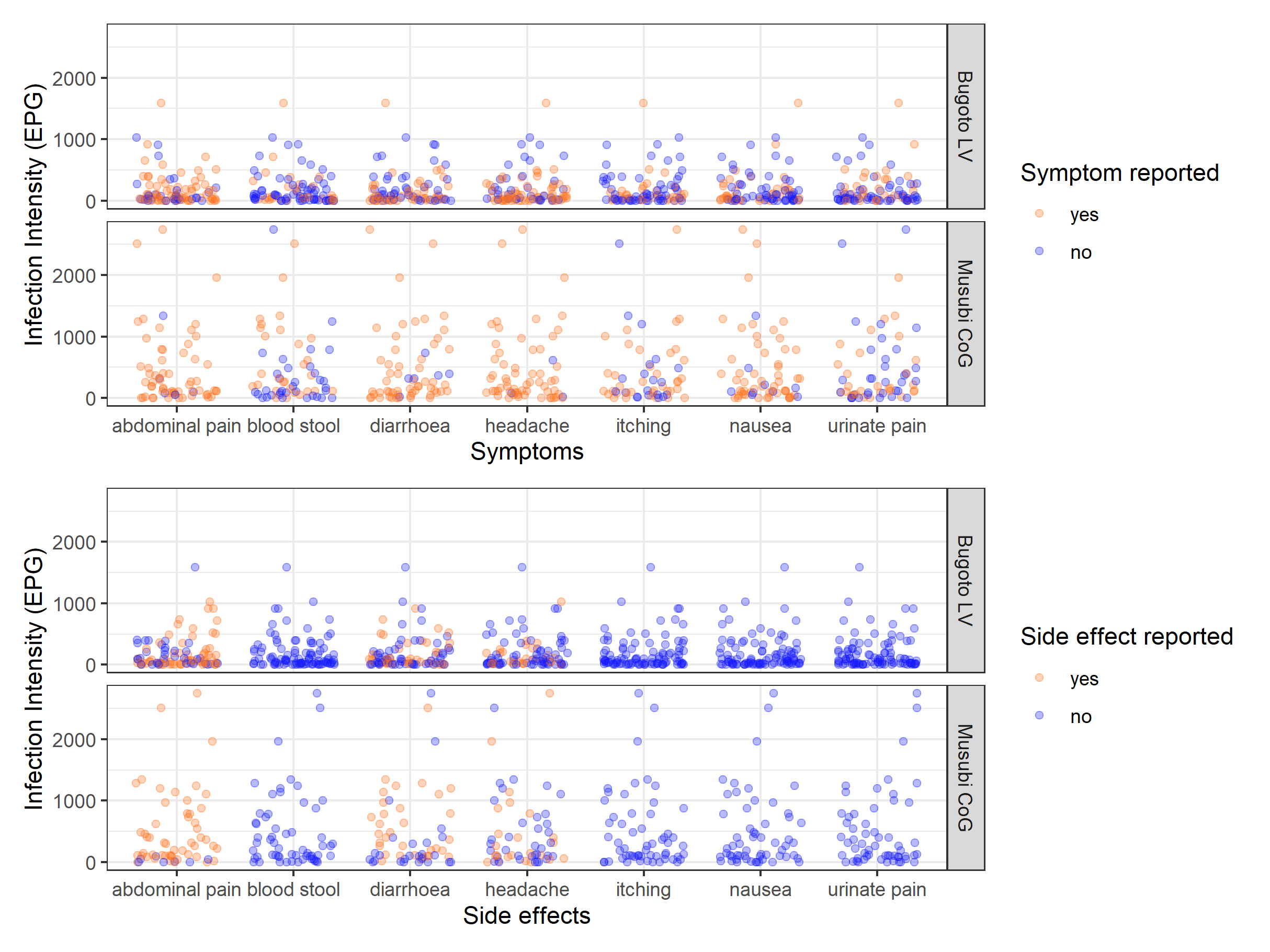

**Figure S1.** The *Schistosoma mansoni* infection intensity of every child in Bugoto Lake View (LV) (top graph in both panel) and Musubi Church of God (CoG) (bottom graph in both panel) primary school who reported (orange), or did not report (blue) pre-treatment symptoms (x-axis of top panel) or post-treatment side effects (x-axis of bottom panel) in 2004. Note the jittering of points to prevent them overlaying one another.

**Musubi COG**

**Symptoms**: Infected students reported pre-treatment symptoms of abdominal pain (79.4%), headache (80.9%), diarrhoea (73.5%), nausea (70.6%). 48.5% of students who got infected reported that they experienced itching, and among infected students, the proportion of students who reported urinating pain (41.2%) and blood stool (39.7%) was comparable to those who did not report them (urinating pain: 42.6%, blood stool: 44.1%) (Figure S2).

**Side effects**: After treatment, there was no student report itching, urinating pain, nausea, and blood stool as side effects. Among infected students, 70.6% of students reported abdominal pain, and the proportions of students who reported headache (30.9%) and diarrhoea (48.5%) as side effects were decreased 50% and 25%, respectively.

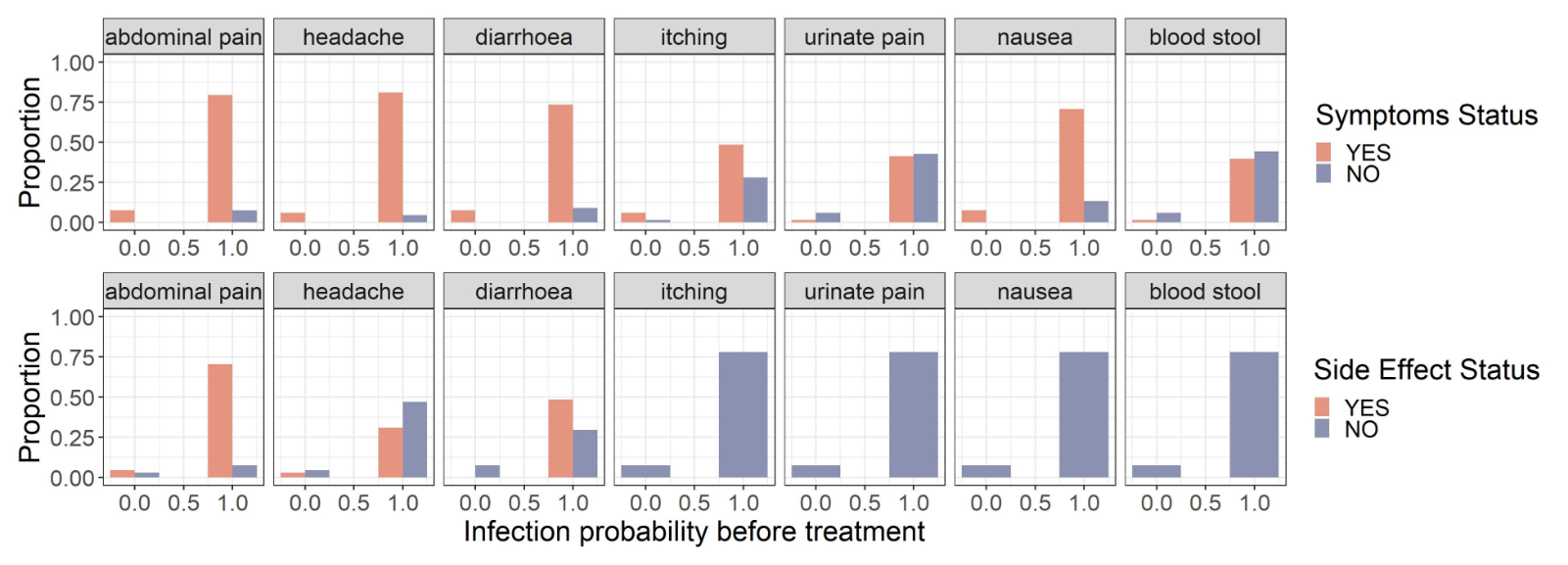

**Figure S2.**	The proportion of Musubi Church of God primary school children who reported (orange), or did not report (purple) pre-treatment symptoms (top row) and post-treatment side effects (bottom row) compared with *Schistosoma mansoni* infection probability.

**Bugoto LV**

**Symptoms**

Less than half of infected students reported headache (50.0%), diarrhoea (42.1%), itching (39.5%), nausea (46.1%), and blood stool (30.3%) as symptoms. However, 64.5% of infected students reported abdominal pain before treatment. (Figure 2). Among infected students, the proportion of students who reported diarrhoea and nausea was similar to those who did not report them (diarrhoea: 47.4%; nausea: 43.4%).

**Side effects**

After treatment, 63.2% of infected students reported abdominal pain as a side effect, and more than half of students who got infected reported that they did not experience headache (65.8%) and diarrhoea (51.3%). There was no student that reported itching, urinating pain, nausea, and blood stool as side effects.

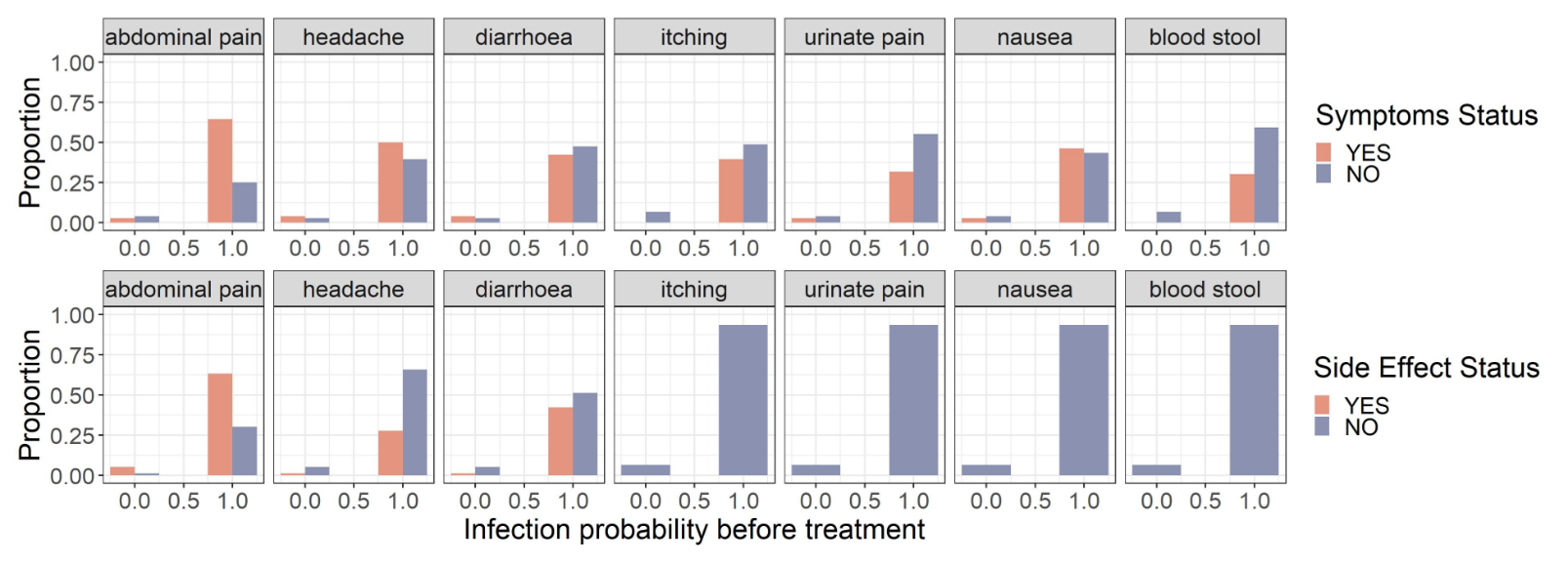

**Figure S3.**	The proportion of Bugoto Lake View primary school children who reported (orange), or did not report (purple) pre-treatment symptoms (top row) and post-treatment side effects (bottom row) compared with *Schistosoma mansoni* infection probability.
